# Comparative performance of the concurrent comparator design with existing vaccine safety surveillance approaches on real-world observational health data

**DOI:** 10.64898/2026.01.25.26344812

**Authors:** Shounak Chattopadhyay, Fan Bu, Martijn J. Schuemie, Jody-Ann McLeggon, Erik Westlund, George Hripcsak, Patrick B. Ryan, Marc A. Suchard

## Abstract

**Background:** It is critical public health concern to identify safety signals originating from wide-scale immunization efforts. Such safety signals may be identified from spontaneous reports and other data sources. Although some work has been done on the best methods for vaccine safety surveillance, there is a scarcity of information on how these perform in analyses of real-world data.

**Methods:** We use four administrative claims databases and one electronic health record (EHR) database to evaluate the operating characteristics of the recently proposed concurrent comparator, self-controlled case series, historical comparator and case-control epidemiological designs for vaccine safety, using negative control outcomes (unrelated to the vaccine), imputed positive control outcomes, and one real-world positive control outcome (myocarditis or pericarditis) for COVID-19. In this evaluation, we consider vaccine exposures for COVID-19, 2017-2018 seasonal influenza, H1N1pdm flu, Human Papillomavirus (HPV), and Varicella-Zoster. The methods are compared based on type 1 error, power of association detection, and proportion of non-finite association estimates produced.

**Results:** All methods exhibit systematic error, leading to type 1 errors that are greater than the nominal (= 0.05) threshold, often by a substantial amount. To restore near-nominal type 1 error, we carry out empirical calibration based on the large set of negative controls. Post-empirical calibration, the self-controlled case series designs had the highest power overall, closely followed by the concurrent comparator designs. However, concurrent comparator analyses often produced a higher proportion of non-finite estimates.

**Conclusion:** Our results indicate that there remains non-negligible systematic error under the concurrent comparator. In terms of statistical performance, the concurrent comparator designs show promising results in some scenarios, regularly outperforming the historical comparator and case-control designs, but often producing non-finite estimates. Future work building on the concurrent comparator design is required to construct more efficient designs with lower systematic error.

## 1 Introduction

Extensive multi-stage randomized clinical trials (RCTs) precede the wide-scale rollout of a vaccine, ensuring its safety. Accurate and efficient reporting of safety signals in the preceding RCTs plays a pivotal role in governing the release and subsequent administration of the vaccine by regulatory agencies. After the wide-scale rollout of a vaccine, it is inevitable that further safety signals are widely reported across different sources. Since RCTs may have a limited number of participants, it is essential to rely in parallel on other forms of information to rapidly identify safety signals from exposure to the vaccine. For instance, it is common to consider observational healthcare data, such as those from electronic health records (EHRs) or administrative claims. The massive sample sizes of observational databases make them particularly suited to detect potential vaccine-induced safety signals which were previously undetected.

There is a rich literature on existing statistical methods for detecting safety signals based on observational health data. Some of the most commonly used epidemiological designs include self-controlled case series (SCCS), self-controlled risk interval (SCRI), cohort studies, and case-control studies. When using such a design, the presence of improperly accounted-for bias can lead to inefficient signal detection or inaccurate estimation of risk corresponding to a potential safety signal. These biases can stem from confounding due to seasonal effects, or individual-level effects such as that of age, sex, or ethnicity. Previous studies (Glanz et al., 2006) indicate substantial bias associated with using case-control designs, while suggesting that SCCS/SCRI designs have inherently lesser bias due to their self-controlled nature which eliminates confounding due to individual differences. However, the risk of confounding remains when using these approaches due to bias from patient-level preferences, such as their choice to get vaccinated or the date of vaccination. Such confounding can lead to further bias when detecting safety signals or obtaining risk estimates.

Motivated by the need to address concerns with residual bias due to preferences of individuals receiving the vaccine, Klein et al. (Klein et al., 2021) recently proposed the concurrent comparator design. The concurrent comparator design first considers a pre-defined risk interval where outcomes will be attributed to the effect of the vaccine; we denote this as the time-at-risk (TAR) interval. It then constructs both the target and comparator cohorts by considering vaccinated patients at-risk and not at-risk, respectively. This reduces confounding arising due to individual-level preferences when receiving the vaccine. The times-at-risk are matched on calendar dates, thus ensuring that time-sensitive trends for both infection and vaccine distribution are similar across both the groups. Furthermore, individuals are also matched based on their demographic characteristics to further reduce confounding. Similar to previous approaches, the concurrent comparator proceeds by constructing a counterfactual to assess the risk of an outcome associated with exposure to a vaccine. However, there exist key differences between different methods based on how the counterfactual is constructed.

Existing state-of-the-art approaches in vaccine safety surveillance are routinely used to detect risks associated with vaccination and identify safety signals during mass vaccination initiatives, such as the one following the recent COVID-19 global pandemic. However, the choice of optimal design for analysis in a given scenario is often unclear. Thus, there is a critical need to assess and compare the efficiency of these methods based on statistical performance metrics. Recent advances in the development of large-scale data analytics software have made it possible to execute such studies (Schuemie et al., 2024). Using open-source software developed within the Observational Health Data Sciences and Informatics (OHDSI) initiative (Hripcsak et al., 2015), Schuemie et al. (2022) carried out an extensive comparative evaluation of the performance of case-control studies, cohort studies, SCCS/SCRI studies, and historical rates-based studies on real-world observational data.

Although the recently proposed concurrent comparator approach has been used to detect safety signals in observational health data (Goddard et al., 2022), there is a research gap in the evaluation of the overall statistical performance characteristics of the concurrent comparator and comparative assessment with existing methods. In this paper, we compare the performance characteristics of the concurrent comparator with the previous approaches considered in Schuemie et al. (2022) on real-world observational data. To the best of our knowledge, our work provides the first such open-source implementation that builds upon the OHDSI tool stack. Our evaluation is based on a large set of real-world negative control outcomes, which are outcomes believed to be unrelated to the effect of the vaccines (Schuemie et al., 2016, 2023; Yang et al., 2023), and positive control outcomes imputed from the original set of negative control outcomes. We carry out a comparative evaluation across five different data sources and eight different vaccine exposures. The data sources include four insurance claims databases and one EHR database. Additionally, we assess the risk of an individual developing myocarditis or pericarditis when exposed to the COVID-19 vaccines. Depending on the results obtained from each method in a particular scenario, such as risk estimation or signal detection, we compare them based on statistical metrics such as type 1 error and power of detection.

## 2 Materials and Methods

### 2.1 Vaccines of Interest

In our analysis, we consider 8 different types of vaccines. Table 1 provides information on the vaccine exposures and the time frame for which they were followed. Along with the specific Fluvirin and Fluzone seasonal flu vaccines, we also consider an analysis that combines all recorded seasonal flu vaccinations during the respective season. We consider 1 dose of the H1N1pdm vaccine and 2 doses of the varicella-zoster (Zoster) and human papillomavirus (HPV) vaccines. For COVID-19, we consider the first doses of two different vaccines. For the Zoster and HPV vaccines requiring 2 doses, we do not stratify our analyses according to dose. Except the historical comparator, we require all other methods to have at least 365 days of continuous observation per subject prior to the date of vaccination.

**Table 1:** Study period for exposures of interest.

| Exposure Name | Start Date | End Date | History Start Date | History End Date |
| --- | --- | --- | --- | --- |
| H1N1pdm vaccination | 01-09-2009 | 31-05-2010 | 01-09-2008 | 31-05-2009 |
| Seasonal flu vaccination (Fluvirin) | 01-09-2017 | 31-05-2018 | 01-09-2016 | 31-05-2017 |
| Seasonal flu vaccination (Fluzone) | 01-09-2017 | 31-05-2018 | 01-09-2016 | 31-05-2017 |
| Seasonal flu vaccination (All) | 01-09-2017 | 31-05-2018 | 01-09-2016 | 31-05-2017 |
| Zoster vaccination (Shingrix) | 01-01-2018 | 31-12-2018 | 01-01-2017 | 31-12-2017 |
| HPV vaccination (Gardasil 9) | 01-01-2018 | 31-12-2018 | 01-01-2017 | 31-12-2017 |
| COVID-19 vaccination (BNT126b2) | 11-12-2020 | 30-06-2021 | 01-01-2019 | 30-06-2019 |
| COVID-19 vaccination (mRNA-1273) | 18-12-2020 | 30-06-2021 | 01-01-2019 | 30-06-2019 |

The study period for each exposure is divided into monthly intervals, with a look at the data being carried out at the end of each month. At each look, we consider data accumulated from the beginning of the study till that month, and define the corresponding duration to be a *time period*. For instance, a time period of 6 months for the BNT126b2 COVID-19 vaccine corresponds to 6 months after the initiation of the study.

### 2.2 Data Sources

The study in this paper uses data from the following five US healthcare data sources:

The IBM MarketScan Commercial Claims and Encounters (CCAE) data source contains adjudicated health insurance claims (e.g. inpatient, outpatient, and outpatient pharmacy) from large employers and health plans who provide private healthcare coverage to employees, their spouses and dependents. Population size `≈` 142 million.

The IBM MarketScan Medicare Supplemental Database (MDCR) contains adjudicated health insurance claims of retirees with primary or Medicare supplemental coverage through privately insured fee-for-service, point-of-service or capitated health plans. Population size `≈` 10 million.

The IBM MarketScan Multi-State Medicaid Database (MDCD) contains adjudicated health insurance claims for Medicaid enrollees from multiple states and includes hospital discharge diagnoses, outpatient diagnoses and procedures, and outpatient pharmacy claims. Population size `≈` 26 million.

The Optum Clinformatics Data Mart - Date of Death (Optum DOD) contains inpatient and outpatient healthcare insurance claims. Population size `≈` 85 million.

The Optum de-identified Electronic Health Record data source (Optum EHR) contains clinical information, prescriptions, lab results, vital signs, body measurements, diagnoses and procedures derived from clinical notes using natural language processing. Population size `≈` 93 million.

### 2.3 Negative Control Outcomes

Our analysis is based on an extensive set of negative control outcomes, which are outcomes with no supporting clinical evidence of being associated with exposure to the vaccine. Ideally, these outcomes would not be reported as safety signals by a vaccine surveillance design. We consider a fixed set of 93 negative controls across the different designs, exposures, and data sources in our analysis.

We initially started with a larger set of potential negative control candidate outcomes, from which the eventual list of 93 negative controls was chosen based on clinical expert opinion. The chosen negative control outcomes match the severity and prevalence of adverse outcomes that could potentially occur from exposure to the vaccine. We chose a large set of negative controls to reliably represent a broad range of diseases or conditions which can capture different population-level systematic error distributions.

In our analysis, we observe a negative control outcome as the first occurrence of the negative control concept or any of its descendants.

### 2.4 Imputed Positive Control Outcomes

Positive controls are outcomes for which there is sufficient clinical evidence to support that they are caused by exposure to vaccines. Ideally, a vaccine surveillance design should be able to flag such outcomes as safety signals as early as possible. However, the use of real-world positive controls can lead to significant issues (Schuemie et al., 2018), as established adverse events due to vaccinations have a rare prevalence among individuals. Even if an effect is established by a design, we cannot ascertain the underlying true risk with certainty. Furthermore, as soon as positive control outcomes are detected as safety signals, immediate regulatory action is immediately taken to mitigate the associated public health risk with exposure to the vaccine. This typically conceals the risks associated with such outcomes in real-world observational data. For these reasons, we do not consider real positive controls when evaluating the statistical performance of the designs based on the metrics described in Section 2.8. However, as described in Section 2.5, we also consider one real-world positive control (the risk of developing myocarditis or pericarditis) from COVID-19 vaccination, and evaluate this risk based each of the designs.

Instead, as in Schuemie et al. (2022), we consider imputed positive controls based on the original set of negative controls, with corresponding effect size estimates generated using a conceptually simple simulation approach. Suppose the estimated effect size corresponding to a negative control outcome is *θ*^^^ with 95% confidence interval [*θ_L_, θ_U_*]. If the desired true effect size for the imputed positive control is *ρ*, we simply multiply this effect size estimate and the 95% confidence interval by *ρ* to obtain the effect size estimate and the 95% confidence interval, respectively, for the imputed positive control. That is, the effect size estimate for the imputed positive control is *ρθ*^^^ with 95% confidence interval [*ρθ_L_, ρθ_U_*]. For illustration, suppose a design produces a risk ratio estimate of 0.8 with 95% confidence interval [0.7, 0.9] for a negative control (having true effect size = 1). Then, for an imputed positive control based on this negative control with true effect size *ρ* = 2, the risk ratio estimate and 95% confidence interval are calculated as 2 × 0.8 = 1.6 and 2 × [0.7, 0.9] = [1.4, 1.8], respectively.

### 2.5 Real-world positive control outcome

Along with the negative controls and imputed positive controls, we also consider the risk of developing myocarditis or pericarditis following the BNT126b2 and mRNA-1273 COVID-19 vaccinations as a real-world positive control outcome. We will evaluate all the designs on this positive control outcome at the end of the study period and compare their results on the basis of effect size estimates and whether a safety signal is declared.

### 2.6 Methods to Evaluate

We evaluate all the methods considering a TAR of 1-28 days after vaccine exposure for all the methods. That is, safety events are attributed to the effect of the vaccine for 1-28 days after vaccination.

#### 2.6.1 Concurrent comparator

Despite adjusting for confounders such as age, season, and gender, there remains a risk of confounding and biased estimation in existing vaccine surveillance approaches due to differences between patients who choose to receive a vaccine or when they choose to obtain the vaccine. The concurrent comparator method aims to control for this bias by predefining a risk interval and designating patients in the exposure target cohort as vaccinated patients who were more recently vaccinated at the time of observation and patients in the control group as vaccinated comparators who had been vaccinated at an earlier time (Klein et al., 2021). By comparing the vaccinated subjects at-risk with vaccinated subjects no longer considered to be at-risk, the concurrent comparator attempts to promote similar healthcare-seeking behavior between the comparison groups. Furthermore, the times-at-risk are matched on calendar dates to ensure that infection rate and other temporal factors are similar between the comparison groups. Figure 1 illustrates the concurrent comparator design with a TAR of 1-28 days. The target TAR is 1-28 days post vaccination, while the comparator control period is 29-56 days post vaccination.

**Figure 1:**
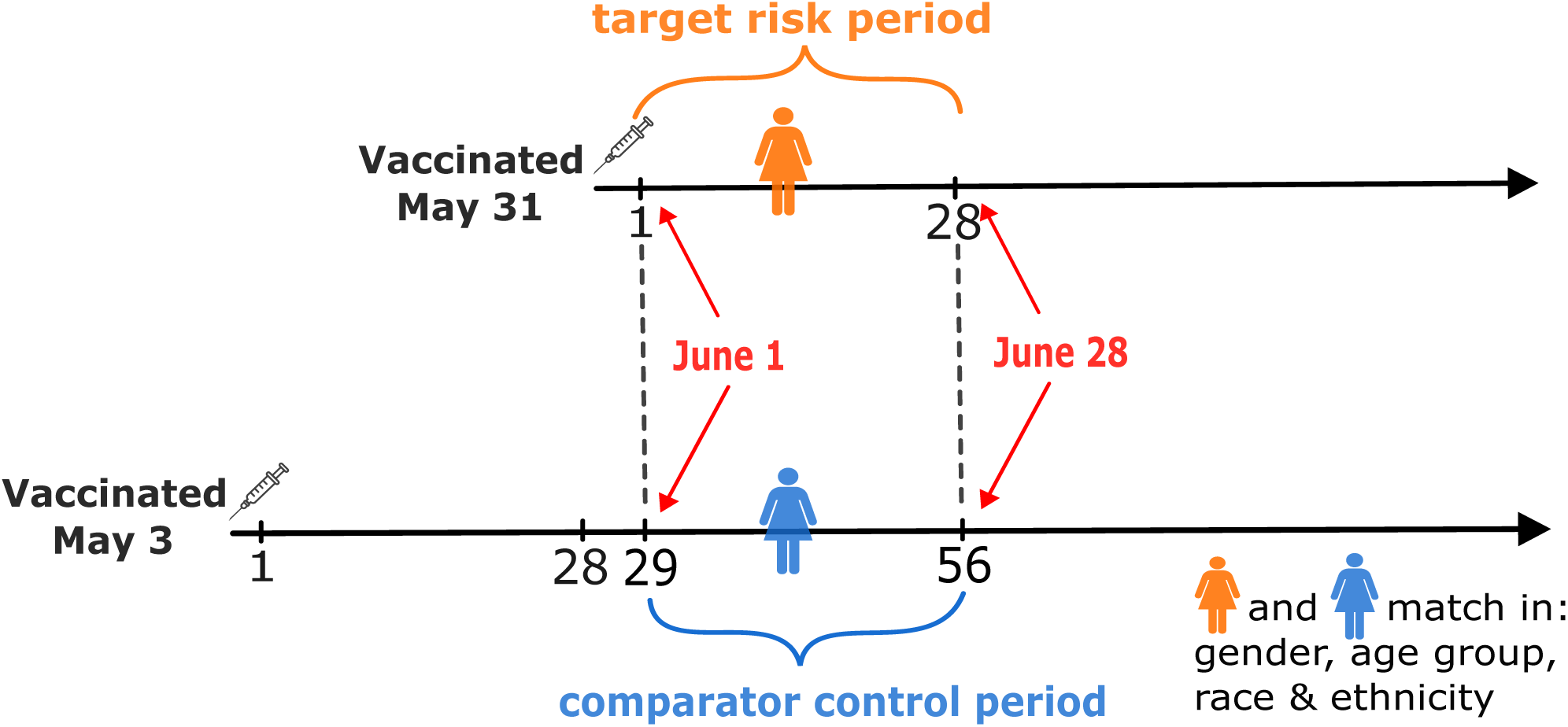
The concurrent comparator approach. The individual (in orange) with an outcome during their target TAR (1-28 days post-vaccination) is matched to another vaccinated individual (in blue) in their comparator control period (29-56 days post-vaccination), based on gender, age group, race, and ethnicity.

#### 2.6.2 Self-Controlled Case Series (SCCS) / Self-Controlled Risk Interval (SCRI)

The SCCS and SCRI designs are self-controlled, comparing the risk-window (the time shortly following the vaccination) to some other time in the same patient’s record. The SCCS design uses all patient time when not at risk as the control time (Whitaker et al., 2006). The SCRI design uses a pre-specified control interval relative to the vaccination date as the control time (Glanz et al., 2006). This unexposed time can be both before or after the time at risk. We will evaluate five variations:

a. A simple SCCS, using all patient time when not at risk as the control time, with the exception of the 30 days prior to vaccination which is excluded from the analysis to avoid bias due to contra-indications.
b. An SCCS adjusting for age and season. Age and season will be modeled to be constant within each calendar month, and vary across months as bicubic splines.
c. A simple SCCS discarding all time prior to vaccination.
d. An SCRI, using a control interval of 43 to 15 days prior to vaccination.
e. An SCRI, using a control interval of 43 to 71 days after vaccination.

#### 2.6.3 Historical Comparator

Traditionally, vaccine surveillance methods compute an expected count based on an incidence rate estimated during some historic time period, for example in the years prior to the initiation of the surveillance study. We will use the historic period indicated in Table 1 and will evaluate the following variations:

a. Unadjusted, entire year: Using a single rate computed across the entire historic year for the entire population.
b. Age and sex adjusted, entire year: Using a rate stratifying by age (in 10 year increments) and sex, computed across the entire historic year. This allows the expected rate to be adjusted for the demographics of the vaccinated.
c. Unadjusted, risk window relative to outpatient visit: Using a single rate computed during the risk window relative to a random outpatient visit in the historic year.
d. Age and sex adjusted, risk window relative to outpatient visit: Using a rate stratifying by age and sex, computed during the risk window relative to a random outpatient visit in the historic year.

Initial results show that this counterfactual approach is sensitive to changes in coding practices. We therefore introduce a study diagnostic: the percent change in overall incidence rate (across the entire population) between the historic and current time period. For each of the four variations listed above, we add a new variation where effect-size estimates are removed if the change in incidence rate is greater than 50%.

#### 2.6.4 Case-control

The case-control design compares cases (those with the outcome) to controls (those that do not have the outcome), and looks back in time for exposures to a vaccine. We will evaluate two variants:

a. Using up to four age and sex matched controls per case. For age we will use a two-year caliper.
b. By sampling controls from the general non-case population, and adjusting for age and sex in the outcome model. The control sample will be four times the number of controls. Age will be modeled as one variable per 5-year age category.

### 2.7 Data characteristics

In Tables 2 and 3, we provide some important data characteristics for the COVID-19 vaccines and the other vaccines, respectively. For both the tables, we provide information on the number of subjects exposed to the vaccine, the duration of exposure aggregated across the exposed subjects, and summary statistics for the negative control outcome counts. For the COVID-19 vaccines, we also provide outcome counts for the positive control (myocarditis/pericarditis).

**Table 2:** Data characteristics for COVID-19 vaccines. “Exposure Subjects” provides the total number of unique vaccinated subjects. “Exposure time (Person-years)” provides the cumulative risk window aggregated over all the vaccinated subjects. “Outcome Counts” provides outcome counts for the realworld positive control outcome (myocarditis/pericarditis), median of negative control outcome counts, and the 25*^th^* and 75*^th^* percentiles of the negative control outcome counts.

|  | Exposure<br>Subjects | Exposure time<br>(Person-years) | Outcome Counts |  |  |
| --- | --- | --- | --- | --- | --- |
|  |  |  | Positive Control<br>(Myocarditis/Pericarditis) | Negative Controls<br>(Median) | Negative Controls<br>(25% - 75%) |
| **COVID-19 vaccination (BNT126b2)** |  |  |  |  |  |
| Optum EHR | 4,573,105 | 648,783 | 382 | 173 | 48 - 528 |
| MDCD | 279,076 | 33,450 | 30 | 12 | 2 - 49 |
| MDCR | 18,496 | 2,529 | 2 | 1 | 0 - 3 |
| Optum DOD | 1,256,748 | 169,306 | 128 | 34 | 5 - 88 |
| CCAE | 2,280,759 | 308,028 | 186 | 42 | 11 - 145 |
| **COVID-19 vaccination (mRNA-1273)** |  |  |  |  |  |
| Optum EHR | 843,717 | 119,018 | 57 | 31 | 7 - 103 |
| MDCD | 213,495 | 26,645 | 23 | 13 | 3 - 42 |
| MDCR | 18,182 | 2,384 | 2 | 1 | 0 - 3 |
| Optum DOD | 793,323 | 107,570 | 59 | 22 | 5 - 62 |
| CCAE | 1,407,524 | 192,862 | 108 | 35 | 8 - 104 |

**Table 3:** Data characteristics for other vaccines, with columns as in Table 2.

|  | Exposure<br>Subjects | Exposure time<br>(person-years) | Outcome Counts |  |
| --- | --- | --- | --- | --- |
|  |  |  | Negative Controls<br>Median, 50% | Negative Controls<br>25% - 75% |
| **Seasonal flu vaccination (all)** |  |  |  |  |
| Optum EHR | 2,536,334 | 190,404 | 100 | 42 - 251 |
| MDCD | 1,237,934 | 94,099 | 30 | 11 - 120 |
| MDCR | 264,636 | 20,093 | 12 | 3 - 34 |
| Optum DOD | 3,399,471 | 259,141 | 142 | 42 - 285 |
| CCAE | 3,516,811 | 266,979 | 66 | 15 - 190 |
| **Seasonal flu vaccination (Fluvirin)** |  |  |  |  |
| Optum EHR | 14,706 | 1,121 | 0 | 0 - 2 |
| MDCD | 15,282 | 1,161 | 0 | 0 - 2 |
| MDCR | 822 | 62 | 0 | 0 - 0 |
| Optum DOD | 189,184 | 14,382 | 4 | 1 - 12 |
| CCAE | 119,186 | 9,029 | 2 | 0 - 6 |

Table 3 (continued).
|  | Exposure Subjects | Exposure time (person-years) | Outcome Counts |  |
| --- | --- | --- | --- | --- |
|  |  |  | Negative Controls | Negative Controls |
|  |  |  | Median, 50% | 25% - 75% |
| Seasonal flu vaccination (Fluzone) |  |  |  |  |
| Optum EHR | 337218 | 23707 | 11 | 4 - 27 |
| MDCD | 3357 | 257 | 0 | 0 - 0 |
| MDCR | 34414 | 2618 | 1 | 0 - 3 |
| Optum DOD | 798816 | 61161 | 30 | 7 - 77 |
| CCAE | 957 | 70 | 0 | 0 - 0 |
| H1N1pdm vaccination |  |  |  |  |
| Optum EHR | 156467 | 11971 | 2 | 0 - 7 |
| MDCD | 206865 | 15458 | 2 | 0 - 6 |
| MDCR | 12913 | 976 | 0 | 0 - 2 |
| Optum DOD | 457565 | 34397 | 8 | 1 - 26 |
| CCAE | 753592 | 56399 | 8 | 1 - 32 |
| Zoster vaccination, first or second dose (Shingrix) |  |  |  |  |
| Optum EHR | 220106 | 20851 | 8 | 3 - 30 |
| MDCD | 11556 | 1241 | 1 | 0 - 2 |
| MDCR | 53384 | 6118 | 2 | 0 - 10 |
| Optum DOD | 232669 | 25637 | 11 | 2 - 29 |
| CCAE | 149219 | 16133 | 5 | 1 - 12 |
| Zoster vaccination, first dose (Shingrix) |  |  |  |  |
| Optum EHR | 219,665 | 16,262 | 7 | 2 - 22 |
| MDCD | 11,407 | 853 | 0 | 0 - 2 |
| MDCR | 52,789 | 3,955 | 1 | 0 - 6 |
| Optum DOD | 229,463 | 17,125 | 8 | 1 - 22 |
| CCAE | 148,190 | 11,012 | 3 | 1 - 7 |
| Zoster vaccination, second dose (Shingrix) |  |  |  |  |
| Optum EHR | 63,464 | 4,589 | 2 | 0 - 6 |
| MDCD | 5,379 | 388 | 0 | 0 - 1 |
| MDCR | 30,218 | 2,163 | 1 | 0 - 3 |
| Optum DOD | 119,556 | 8,513 | 3 | 1 - 9 |
| CCAE | 72,063 | 5,121 | 1 | 0 - 4 |
| HPV vaccination, first or second dose (Gardasil 9) |  |  |  |  |
| Optum EHR | 234,518 | 19,321 | 2 | 0 - 8 |
| MDCD | 237,455 | 18,846 | 1 | 0 - 5 |
| MDCR | 0 | 0 | 0 | 0 - 0 |
| Optum DOD | 174,692 | 14,457 | 1 | 0 - 6 |
| CCAE | 378,052 | 31,712 | 1 | 0 - 8 |
| HPV vaccination, first dose (Gardasil 9) |  |  |  |  |
| Optum EHR | 233,985 | 17,313 | 1 | 0 - 6 |
| MDCD | 236,683 | 17,779 | 1 | 0 - 5 |
| MDCR | 0 | 0 | 0 | 0 - 0 |
| Optum DOD | 173,228 | 12,948 | 0 | 0 - 6 |
| CCAE | 376,341 | 28,222 | 1 | 0 - 8 |
| HPV vaccination, second dose (Gardasil 9) |  |  |  |  |
| Optum EHR | 28,336 | 2,007 | 0 | 0 - 1 |
| MDCD | 15,065 | 1,067 | 0 | 0 - 0 |
| MDCR | 0 | 0 | 0 | 0 - 0 |
| Optum DOD | 21,377 | 1,510 | 0 | 0 - 0 |
| CCAE | 49,283 | 3,489 | 0 | 0 - 1 |

### 2.8 Performance Metrics

The use of negative controls and imputed positive controls allows us to evaluate the competing methods based on the following criteria:

- **Type 1 Error:** The probability of a negative control outcome being declared as a safety signal. In our study, we estimate type 1 error of a design as the proportion of negative controls flagged as safety signals.
- **Power of Detection:** Also known as sensitivity, it is the probability of a positive control outcome being declared as a safety signal. In our study, we estimate power of a design by the proportion of imputed positive controls flagged as a safety signal and stratify the results by their true effect size.
- **Proportion of non-finite estimates:** The proportion of negative controls for which the method was unable to produce a finite effect size estimate and associated standard error.

Since this is a sequential setup, we will evaluate type 1 error and power of detection for a design across time periods during the respective study period.

### 2.9 Sequential Decision Rule

When declaring whether an outcome is a safety signal, we are interested in the following one-sided hypothesis testing problem:

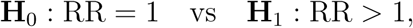

where RR denotes the outcome-specific risk ratio from exposure to a particular vaccine. For negative control outcomes, the true RR equals 1, while for the imputed positive control outcomes, the true RR equals the true effect size used for imputation.

To adjust for multiple testing of the same hypothesis across different time periods, we use the sequential MaxSPRT decision rule (Kulldorff et al., 2011) for declaring an outcome as a safety signal. For each negative control or imputed positive control outcome and a given time period, we first compute a critical value and declare an outcome to be a safety signal if the corresponding one-sided log-likelihood ratio (LLR) exceeds the previously computed critical value. If an outcome is declared a safety signal at a particular time period, it remains declared as a safety signal for all subsequent time periods.

### 2.10 Systematic Error and Empirical Calibration

For a particular design, its type 1 error may deviate substantially from the nominal value (taken to be 0.05) due to unadjusted systematic error. The nature of systematic error may include any form of inherent unadjusted bias when using a design, such as unmeasured confounding, measurement error, and selection bias. Without restoring nominal or near-nominal type 1 errors of the methods, it is inappropriate to compare their performances on the basis of power of detection. This is because one could arbitrarily modify the threshold for declaring a safety signal to achieve any desired power if the type 1 error is allowed to be unrestricted. Thus, to restore the type 1 errors of the methods to (near) nominal values, we employ the empirical calibration approach in Schuemie et al. (2014). Empirical calibration uses effect size estimates and standard errors corresponding to the negative control outcomes to first fit a systematic error distribution (assumed to be Gaussian), and then use the fitted systematic error distribution to calibrate effect size estimates, such as risk ratios, confidence intervals, p-values, and LLRs. This calibration ensures that the approaches have approximately nominal frequentist properties under the null hypothesis.

Although not always, the effect of empirical calibration typically increases p-values and decreases LLRs. This effect decreases both type 1 error and power of detection, regardless of whether or not a sequential decision rule like MaxSPRT is employed to declare safety signals. The amount of discrepancy of these quantities with their uncalibrated values usually increases with increasing systematic error of the design. When calibrating the effect size estimate for a negative control outcome, we proceed with a leave-one-out approach, calibrating corresponding effect size estimates with the systematic error distribution obtained using all the other negative control outcomes.

## 3 Results

### 3.1 Preliminaries

As it is impractical to present results for all of the variants for a particular design due to the large number of possible analyses, we choose to focus on one specific variant for each design which are kept fixed across all the analyses, with all the variants having a TAR of 1-28 days. For the casecontrol approach, we use age and sex matched controls. For the historical comparator approach, we adjust by age and sex and use the TAR after historic visit. For the SCCS, we adjust by age and season, excluding the pre-vaccination window. Overall, the chosen variants in this work were found to perform the best overall across a wide variety of scenarios in Schuemie et al. (2022), which compared numerous existing variants of the SCCS, case-control, and historical designs.

We first consider the systematic error, type 1 error (before and after empirical calibration), and power of detection of the methods. For this, we will consider the Optum EHR data source and exposure to both the BNT126b2 and mRNA-1273 COVID-19 vaccines, all seasonal flu vaccines, H1N1pdm vaccine, Zoster vaccine, and HPV vaccine. We also compare the methods based on the proportion of non-finite estimates produced in both the Optum EHR and MDCD data sources, considering the H1N1pdm vaccination. Next, we consider the variation of these metrics across different data sources. For this, we consider the BNT126b2 and mRNA-1273 COVID-19 vaccines based on the Optum EHR and MDCD data sources. Finally, we estimate the risk of an individual developing myocarditis or pericarditis from exposure to the BNT126b2 and mRNA-1273 COVID-19 vaccines based on the Optum EHR data source, and compare the estimates across the methods.

### 3.2 Systematic Error

In Figure 2, we plot the fitted density functions corresponding to the systematic error distributions of the methods. Ideally, systematic error densities should be centered at true risk ratio = 1 with 0 variance, which is not the case in Figure 2. For instance, the systematic error density for the casecontrol approach can be noticeably biased away from the origin, such as for the seasonal flu and H1N1pdm vaccines, while the density for the historical comparator approach has a high variance across exposures. Additionally, the error distributions often vary considerably across exposures. For instance, the density for SCCS has a much higher variance for the HPV vaccine compared to the H1N1pdm vaccine. In general, we observe the systematic error densities for the self-control case series and concurrent comparator designs to be centered closer to a risk ratio of 1.

**Figure 2:**
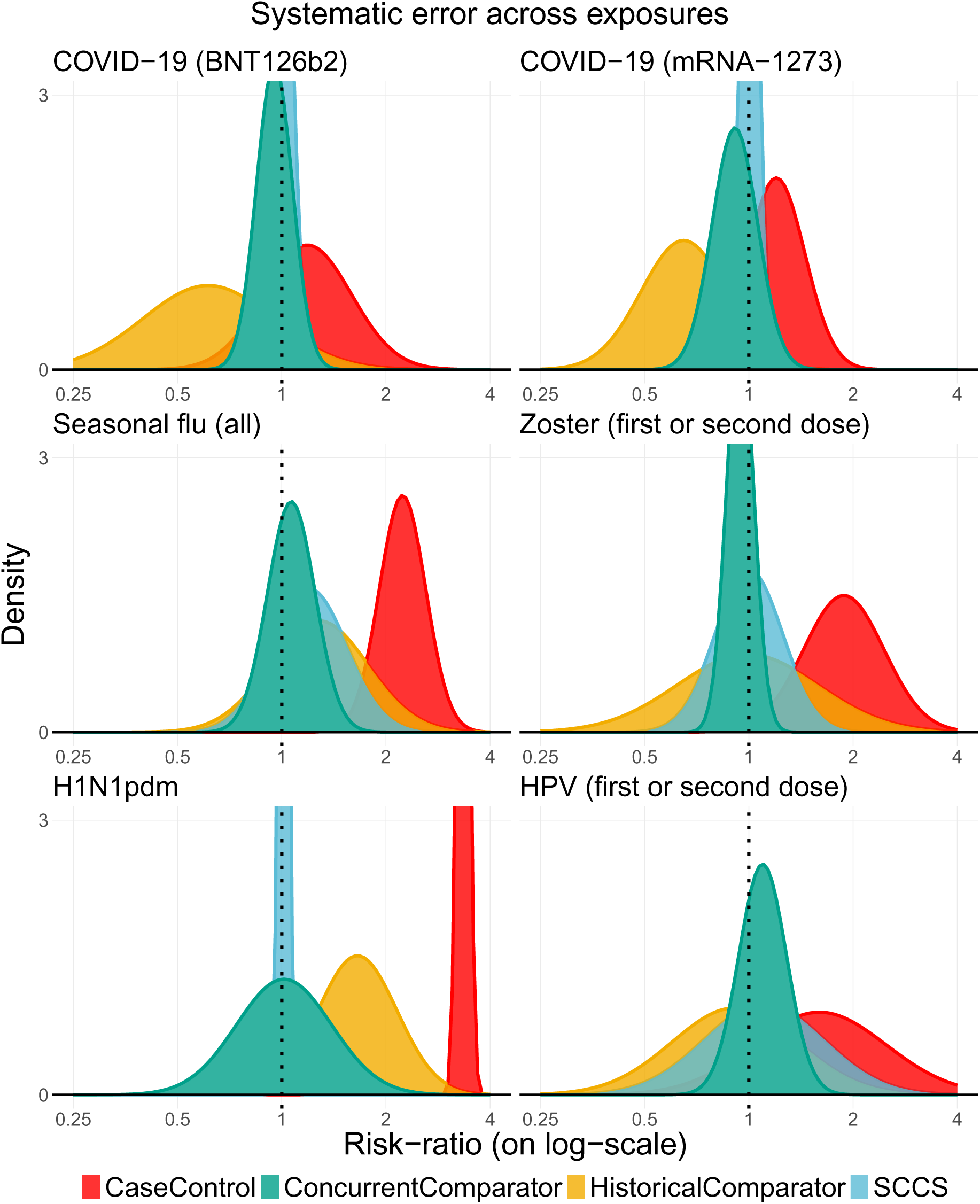
Systematic error for the methods across different exposures, considering the Optum EHR data source.

### 3.3 Type 1 Error and Power of Detection

In Figure 3, we plot the type 1 errors of the designs across time before empirical calibration and stratify the results across exposures. Due to the varying degrees of systematic error, the uncalibrated type 1 errors across observed time periods often differ substantially from the desired nominal value (taken to be 0.05). The concurrent comparator and SCCS approaches have type 1 errors close to the nominal value for some exposures, such as the H1N1pdm and HPV vaccines, while the case-control and historical comparator approaches often have type 1 errors far exceeding the nominal threshold.

**Figure 3:**
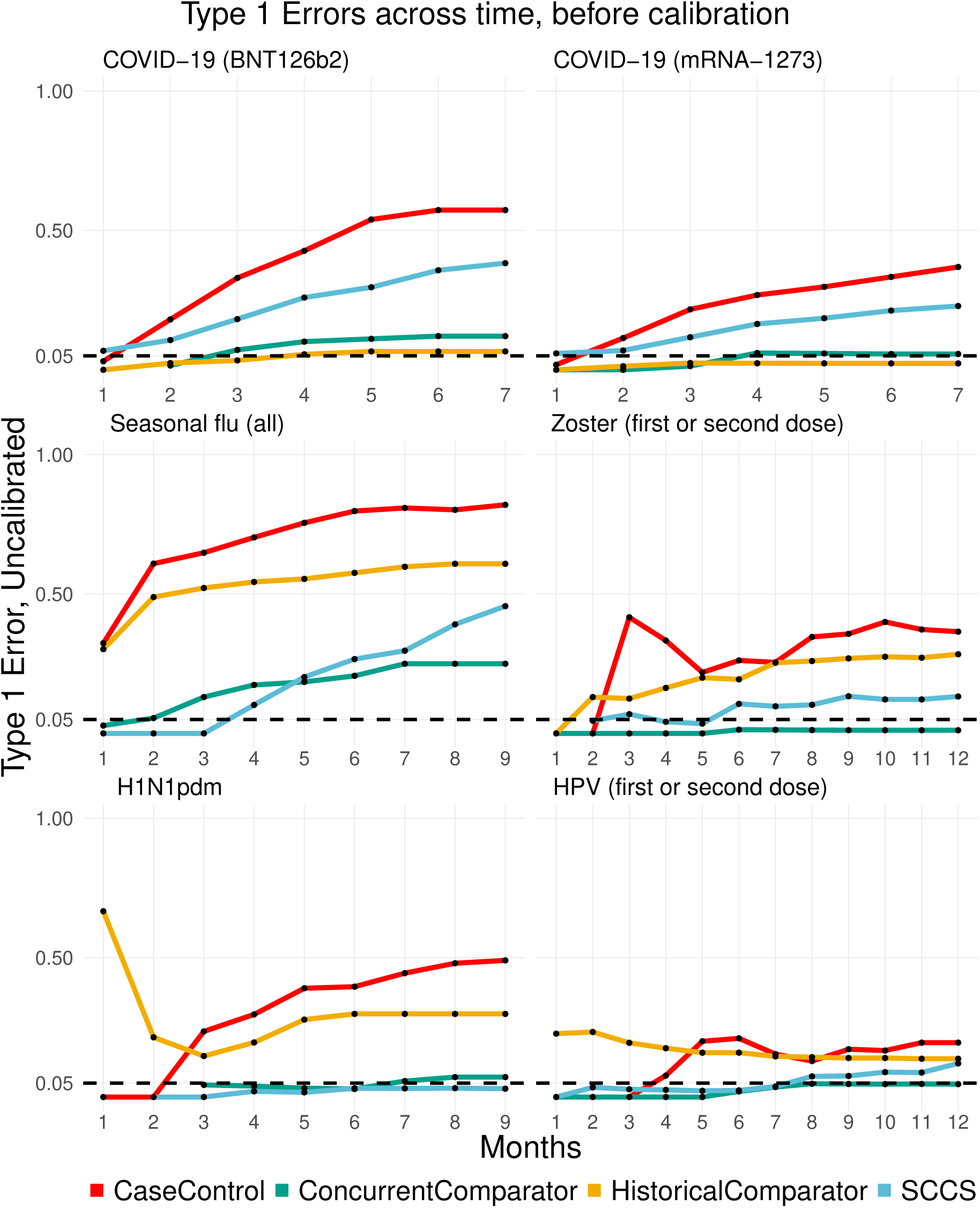
Uncalibrated Type 1 errors of methods as a function of time, across different exposures for the Optum EHR data source.

Next, we proceed to compare type 1 error and power of detection between methods across the observed time periods, after empirical calibration. When evaluating power, we let the true effect size *ρ* for the imputed positive controls be *ρ* = 2. Results for *ρ* ∈ {1.5, 4} are available in the Supplementary Material. Figures 4 and 5 compare the type 1 error and power between the methods across time, respectively, after empirical calibration, with the results stratified across exposures as before. The effect of empirical calibration restores near-nominal type 1 errors for the methods in most of the cases. However, there remain cases with type 1 errors greater than the nominal threshold post-calibration, such as with SCCS for both COVID-19 vaccines, while the concurrent comparator exceeds the threshold for the seasonal flu vaccine. The case-control design often exhibits calibrated type 1 errors noticeably greater than the nominal threshold, such as for the Zoster and HPV vaccines. In terms of power of detection across time, the SCCS performs the best for the most number of cases, with the concurrent comparator close to the SCCS. In terms of timeliness in detecting a safety signal, the concurrent comparator outperforms its competitors in some scenarios, such as for the seasonal flu vaccine. For most of the cases, the historical comparator has a greatly reduced power after calibration, leading to poor performance.

**Figure 4:**
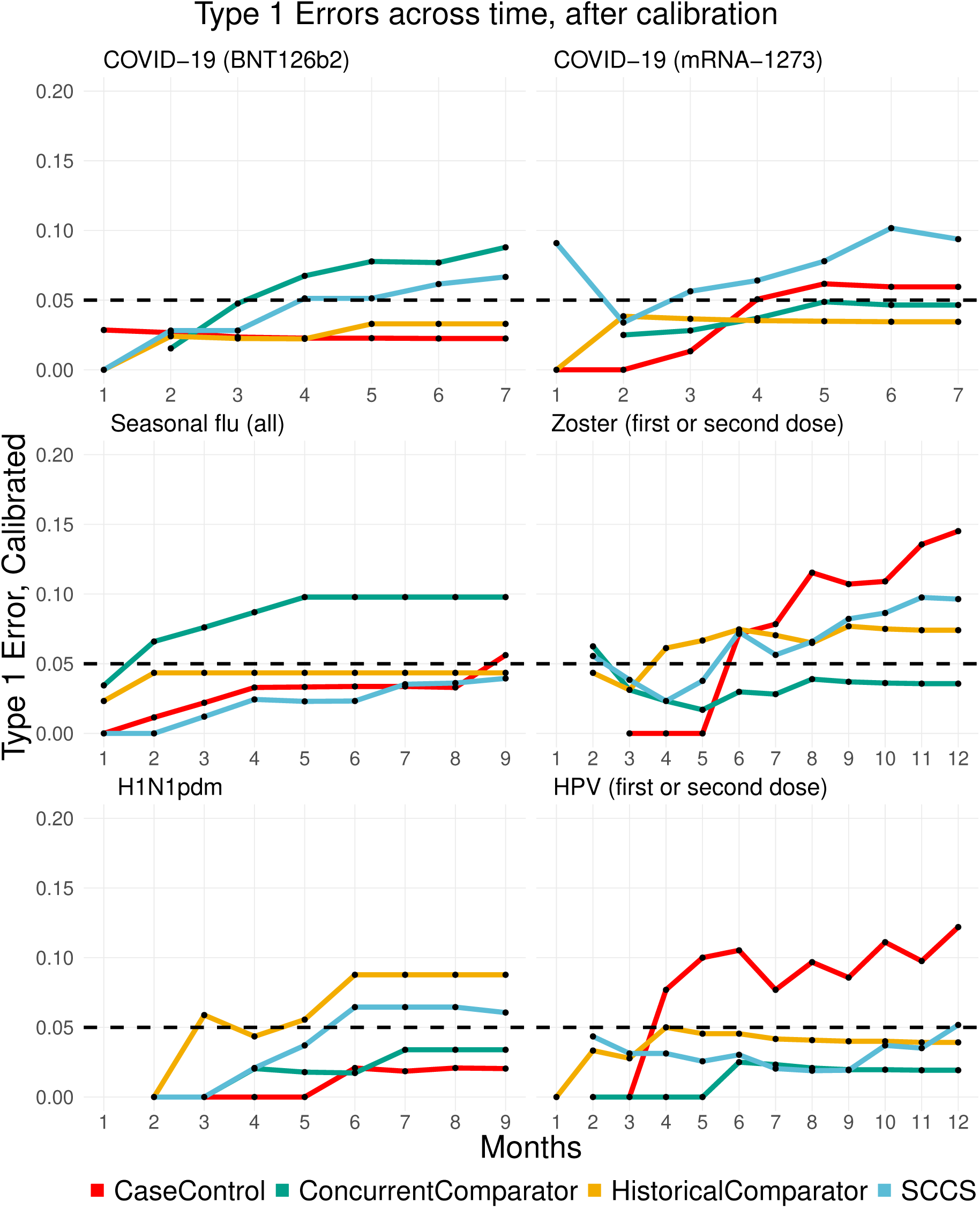
Calibrated type 1 errors of methods as a function of time, across different exposures for the Optum EHR data source.

**Figure 5:**
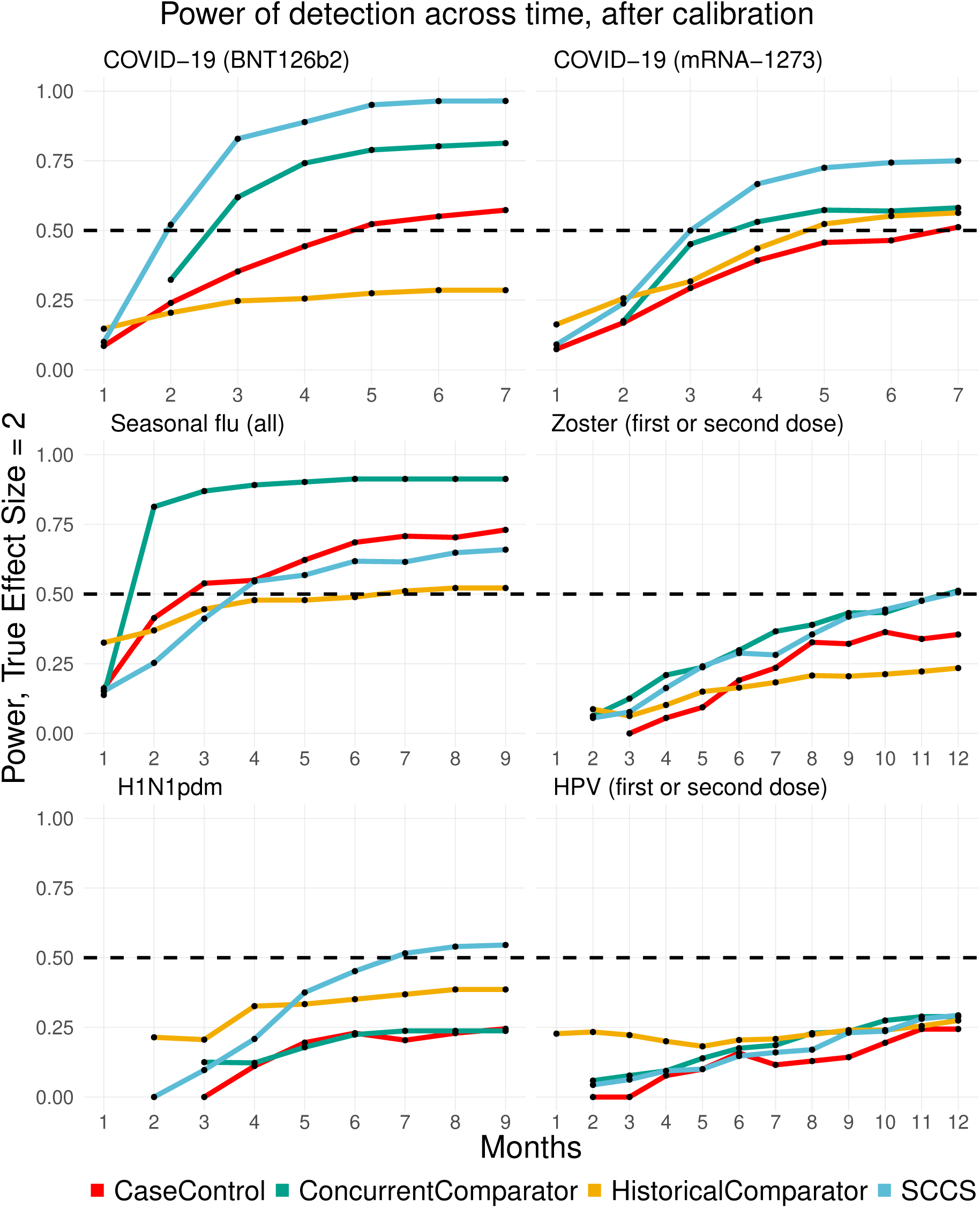
Power of detection for the methods as a function of time, across different exposures for the Optum EHR data source. We assume the true risk ratio = 2.

### 3.4 Proportion of non-finite estimates

In certain scenarios, a design may not produce a finite effect size estimate when evaluating the risk of developing a particular outcome from exposure to the vaccine. There could be various reasons for this, such as there being no outcomes in either the target or comparator cohort, or due to artifacts of stratification when constructing the cohorts. In Table 4, we demonstrate the proportion of nonfinite estimates produced by each design with the H1N1pdm vaccine, considering the entire data observed till the end of study period for the Optum EHR and MDCD data sources. The concurrent comparator has a higher proportion of non-finite estimates produced when compared to all the other methods, possibly due to the intensive stratification when constructing the counterfactuals.

**Table 4:**
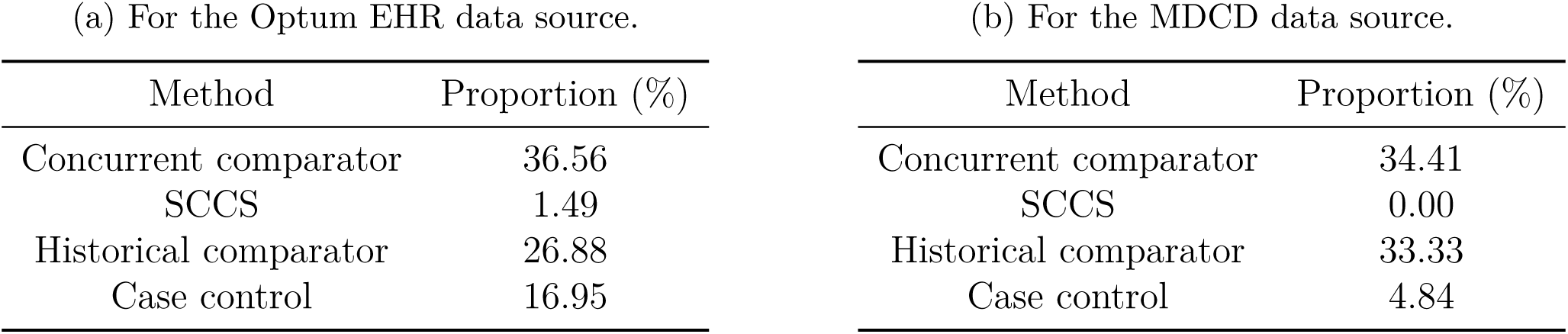
Proportion of non-finite estimates produced by each method for the H1N1pdm vaccine, considering the Optum EHR and MDCD data sources.

| (a) For the Optum EHR data source. |  | (b) For the MDCD data source. |  |
| --- | --- | --- | --- |
| Method | Proportion (%) | Method | Proportion (%) |
| Concurrent comparator | 36.56 | Concurrent comparator | 34.41 |
| SCCS | 1.49 | SCCS | 0.00 |
| Historical comparator | 26.88 | Historical comparator | 33.33 |
| Case control | 16.95 | Case control | 4.84 |

### 3.5 Variation across Data Sources

The performance metrics may often vary for different data sources. We first consider the systematic error densities, with the corresponding plots available in Figure 6. The density for the concurrent comparator has lesser variance for the COVID-19 (BNT126b2) vaccine in the larger Optum EHR data source compared to the smaller MDCD data source. Interestingly, no such difference is noticeable for the COVID-19 (mRNA-1273) vaccine. Across all the four cases, the densities for the historical comparator and case-control approaches are biased away from the null, with the density for the historical comparator having a large variance.

**Figure 6:**
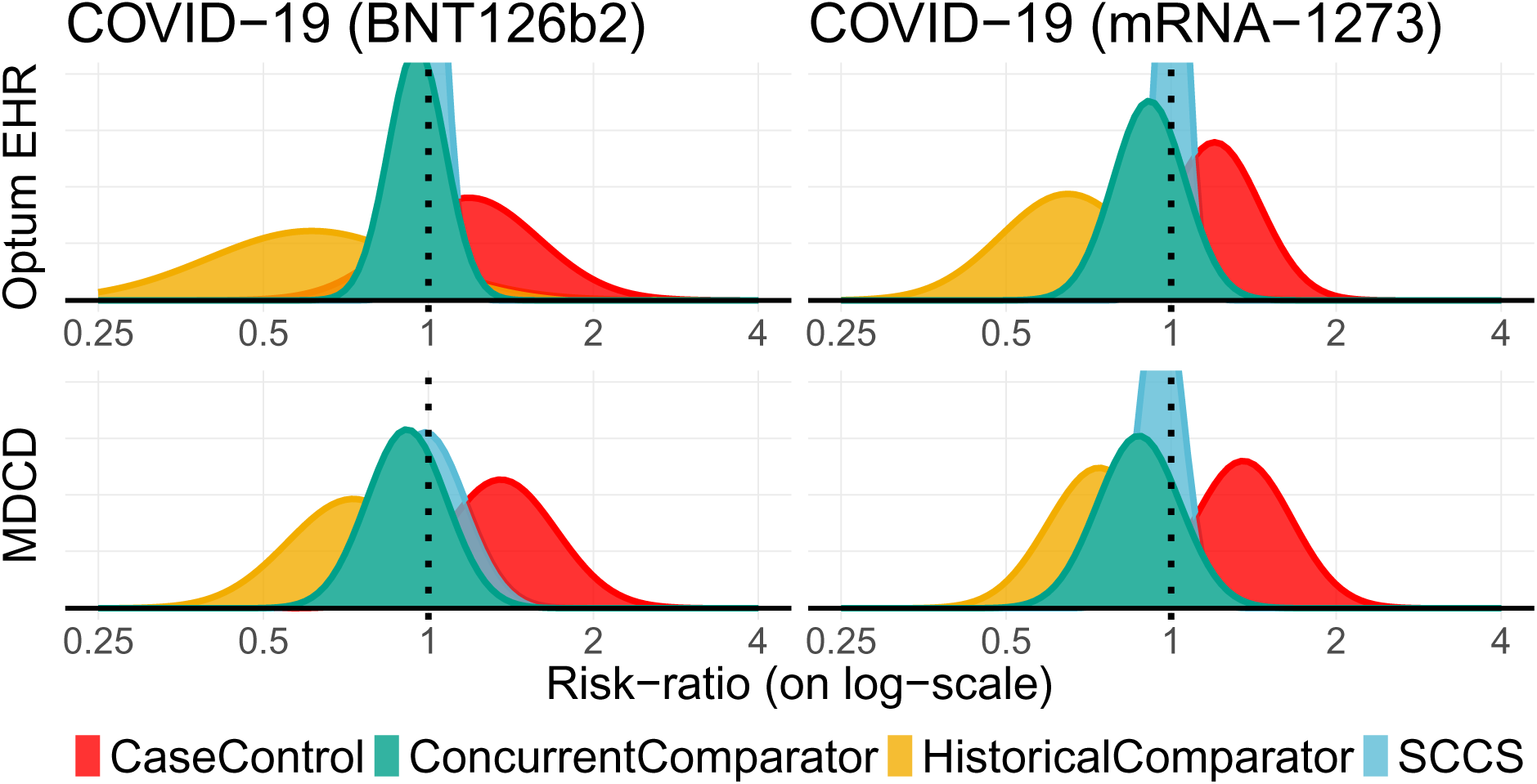
Systematic error for the methods, stratified across the two Covid-19 vaccines and the Optum EHR and MDCD data sources.

After empirical calibration to restore type 1 error, there are some differences in the power of detection for the designs across data sources. The corresponding plots for type 1 error and power are available in Figures 7 and 8, respectively. The concurrent comparator has substantially better power for the larger Optum EHR data source than the MDCD data source for the BNT126b2 COVID-19 vaccine. For both the vaccines, the concurrent comparator has a power greater than 0.50 after the end of the study period in the Optum EHR data source, while falling short of this mark for the MDCD data source. Across all the four cases considered, SCCS performs the best, but often at the cost of increased type 1 error beyond the nominal threshold.

**Figure 7:**
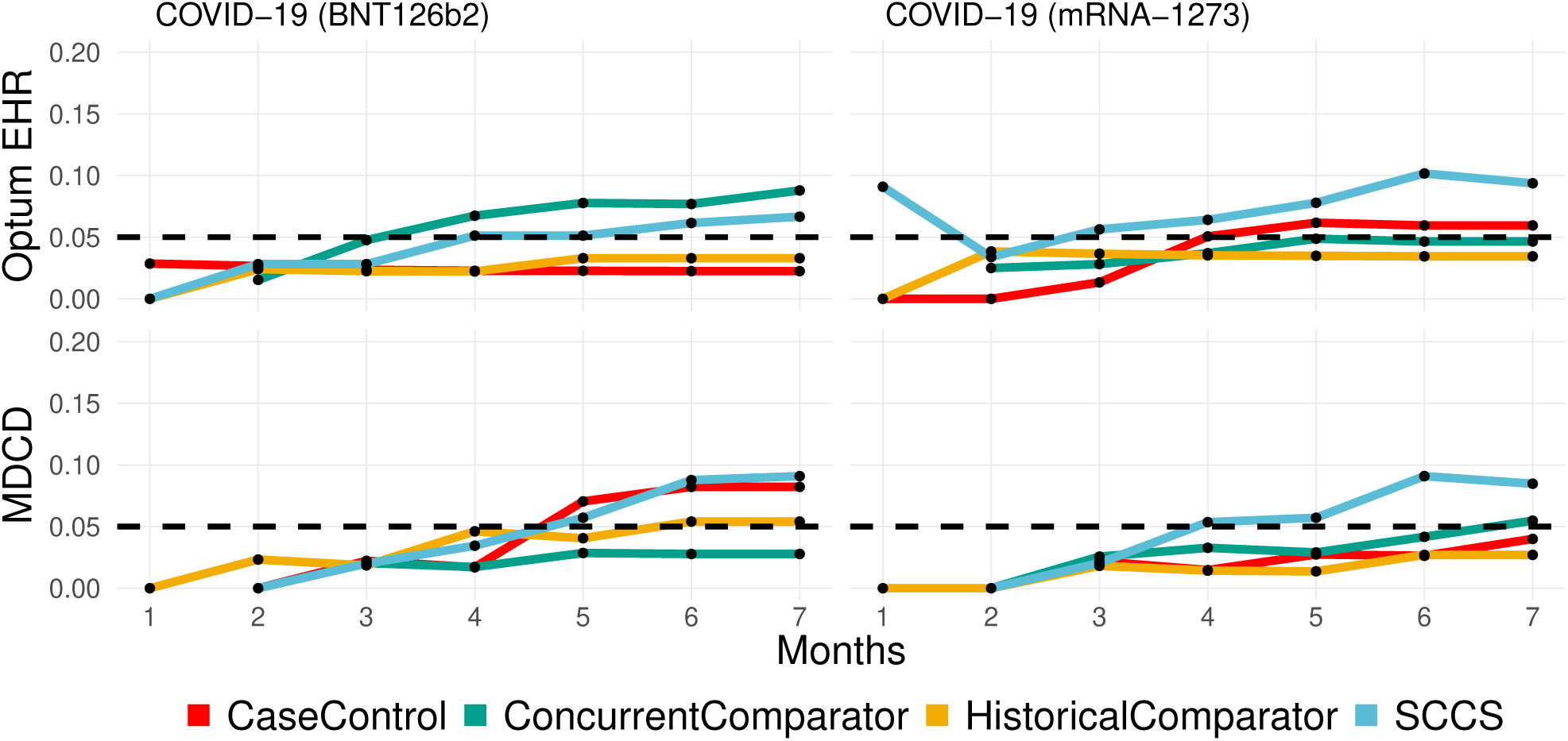
Calibrated type 1 error for the methods as a function of time, stratified across the two Covid-19 vaccine exposures and the Optum EHR and MDCD data sources.

**Figure 8:**
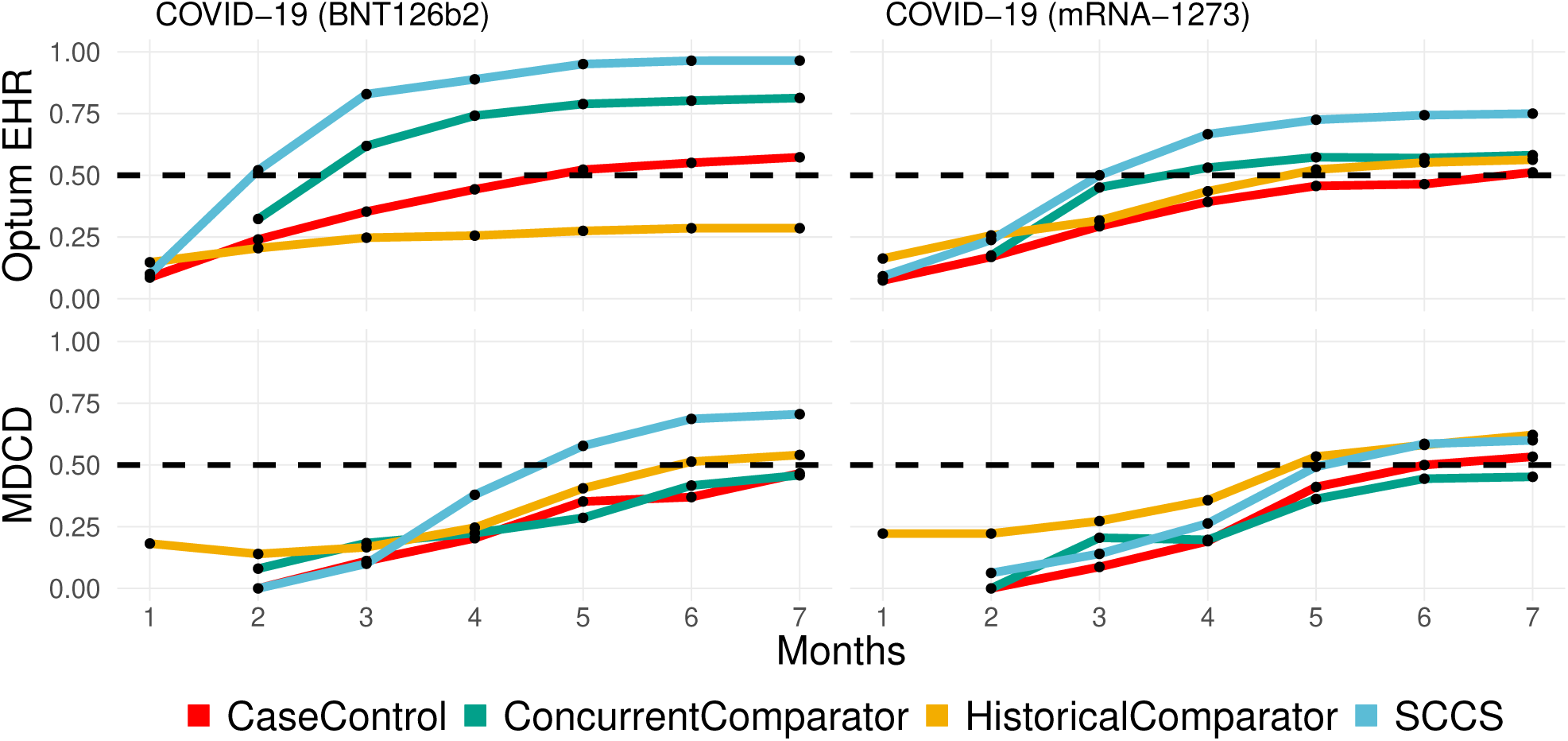
Calibrated power of detection for the methods as a function of time, stratified across the two Covid-19 vaccine exposures and the Optum EHR and MDCD data sources.

### 3.6 Risk of Myocarditis or Pericarditis

We now consider estimating the risk of myocarditis or pericarditis following exposure to the two COVID-19 vaccines. We report our findings at the end of the study period in Table 5, with all effect sizes estimates reported after empirical calibration. None of the methods declare a safety signal for this outcome at a significance level of 0.05, across the two different vaccine types.

**Table 5:**
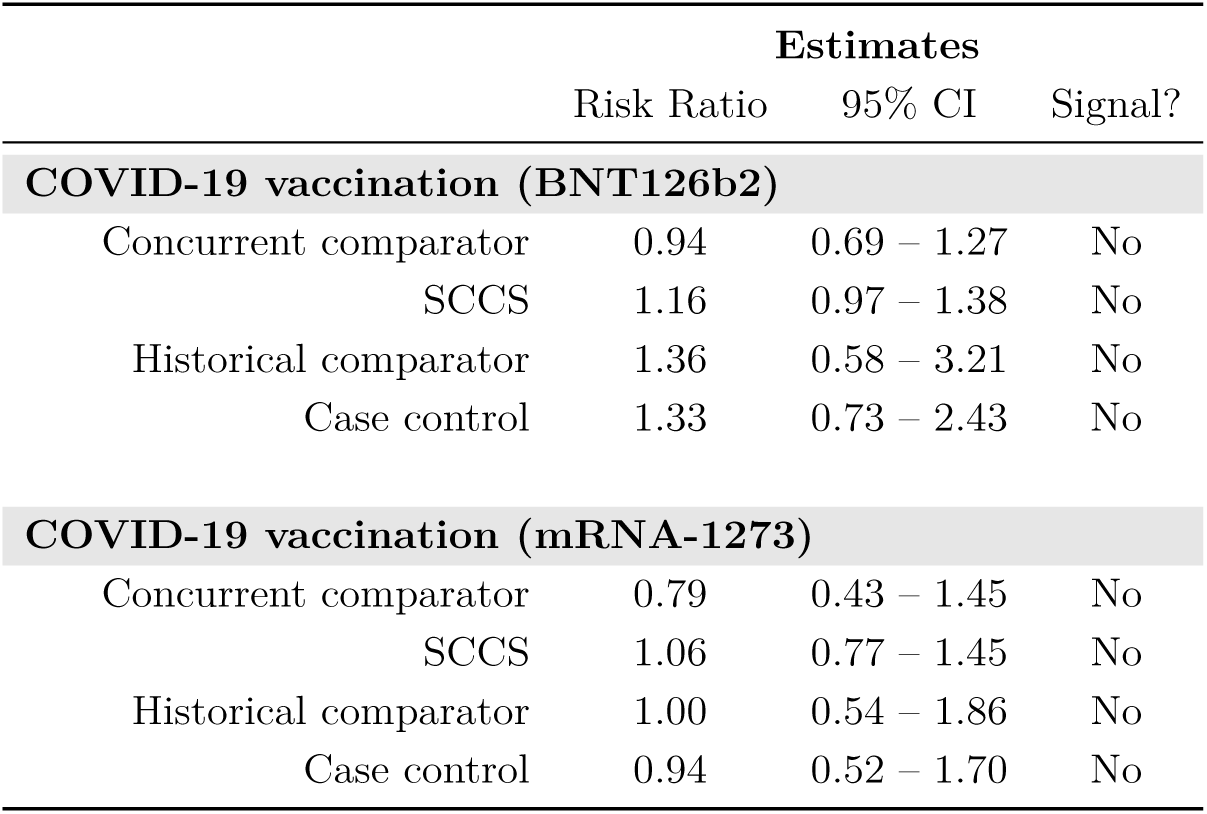
Risk of developing myocarditis or pericarditis following vaccination with COVID-19 BNT126b2 and mRNA-1273 vaccines for the Optum EHR data source. Each table shows the estimated risk ratio with the lower and upper bounds of the 95% confidence interval after empirical calibration, and whether or not a signal was declared.

## 4 Discussion

### 4.1 Strengths and Limitations

There are certain key strengths of our study. Firstly, our study quantifies the amount of systematic error present when implementing a design through the use of negative control outcomes, subsequently adjusting the original estimates via empirical calibration to provide more reliable evidence. Secondly, our study is based on entirely open-source software, with the study protocol available before initiation of the analyses. Thirdly, the protocol clearly mentions the pre-specified selection of covariates, thus avoiding investigator-specific biases in variable selection. Fourthly, the use of multiple data sources, exposures, and outcomes adds credibility to the study and supports generalizability to other settings. Lastly, the dissemination of the obtained results do not depend on the estimated effects, thus avoiding publication bias.

However, there remain certain limitations to our study. Firstly, even though multiple potential confounders were considered when implementing the designs, there could be residual bias due to unmeasured or misspecified confounders. The use of negative controls does alleviate this issue somewhat by allowing us to quantify this residual bias. Secondly, in all the designs, we assume the relative risk from vaccination is not time-varying, which could be restrictive. Thirdly, misclassification of study variables (such as treatments, covariates, and outcomes) is unavoidable when considering observational health data. We do not expect differential misclassification, so bias will most likely be towards the null. Finally, the EHRs used in our study may miss care episodes for patients due to care outside the respective health systems. As before, we expect the resulting bias to be most likely towards the null.

### 4.2 Conclusion

The concurrent comparator design was originally proposed with the intention of reducing inherent biases present in existing vaccine surveillance designs due to patient-level preferences. Instead of ignoring the possibility of further systematic error when implementing the concurrent comparator, the use of negative controls allows us to quantify the amount of systematic error to produce more reliable effect size estimates via empirical calibration. In general, our results indicate a lower degree of systematic bias associated with the concurrent comparator design compared to existing approaches. However, this bias is not exactly zero, which often leads to higher type 1 errors than the allowed nominal threshold.

Overall, in terms of power of detection, the SCCS analyses perform the best for the most number of cases. However, the concurrent comparator is close to the SCCS design for most cases across different exposures, particularly for the larger data sources, while having a generally lower degree of systematic error. In a few cases, the concurrent comparator design outperforms other approaches when detecting safety signals, such as for the seasonal flu vaccine. However, due to its relatively high proportion of non-finite estimates produced, estimates of power and type 1 error are more unreliable compared to other methods. Furthermore, we also found the performance of the concurrent comparator to diminish for smaller data sources, perhaps due to its intensive stratification requiring greater number of subjects to yield reliable estimates.

## Author Contributions

All the authors contributed to the formulation and design of the study. SC, MJS, and MAS implemented the safety surveillance epidemiological designs and executed them across the data sources and exposures. SC, FB, GH, PBR, MJS, and MAS performed the statistical analysis. SC wrote the first draft of the manuscript. All authors were involved in revision of the manuscript, and subsequently read and approved the submitted version.

## Supplementary Material

The Supplementary Material contains plots of the uncalibrated and calibrated type 1 errors, power of detection, systematic error densities, proportion of non-finite estimates, and effect size estimates (with 95% confidence intervals) regarding the risk of myocarditis/pericarditis for the MDCR, MDCD, Optum DOD, and CCAE data sources.

## Data Availability Statement

The patient-level data used for these analyses is not available for public use due to information governance restrictions. The study protocol is available at https://ohdsi-studies.github.io/Eumaeus/Protocol.html, with the source code publicly available at https://github.com/ohdsi-studies/Eumaeus. The authors encourage public contribution and review of the source code, which are free to re-use for future research.

## Ethics Statement

Since this work does not involve human subjects research, the use of the IBM Marketscan and Optum data sources was determined to be exempt from approval by the New England Institution Review Board (IRB) after review. The project does, however, use de-identified human data collected during routine healthcare provision. All data partners executing the research project within their data sources will have received IRB approval or waiver for participation in accordance to their institutional governance prior to execution. The research project executes across a federated and distributed data network, where analysis code is sent to participating data partners and only aggregate summary statistics are returned, with no sharing of patient-level data between organizations.

