## Supplementary Material for "Comparative performance of the concurrent comparator design with existing vaccine safety surveillance approaches on real-world observational health data"

### 1 Systematic error distributions

#### 1.1 Result for the Optum DOD data source

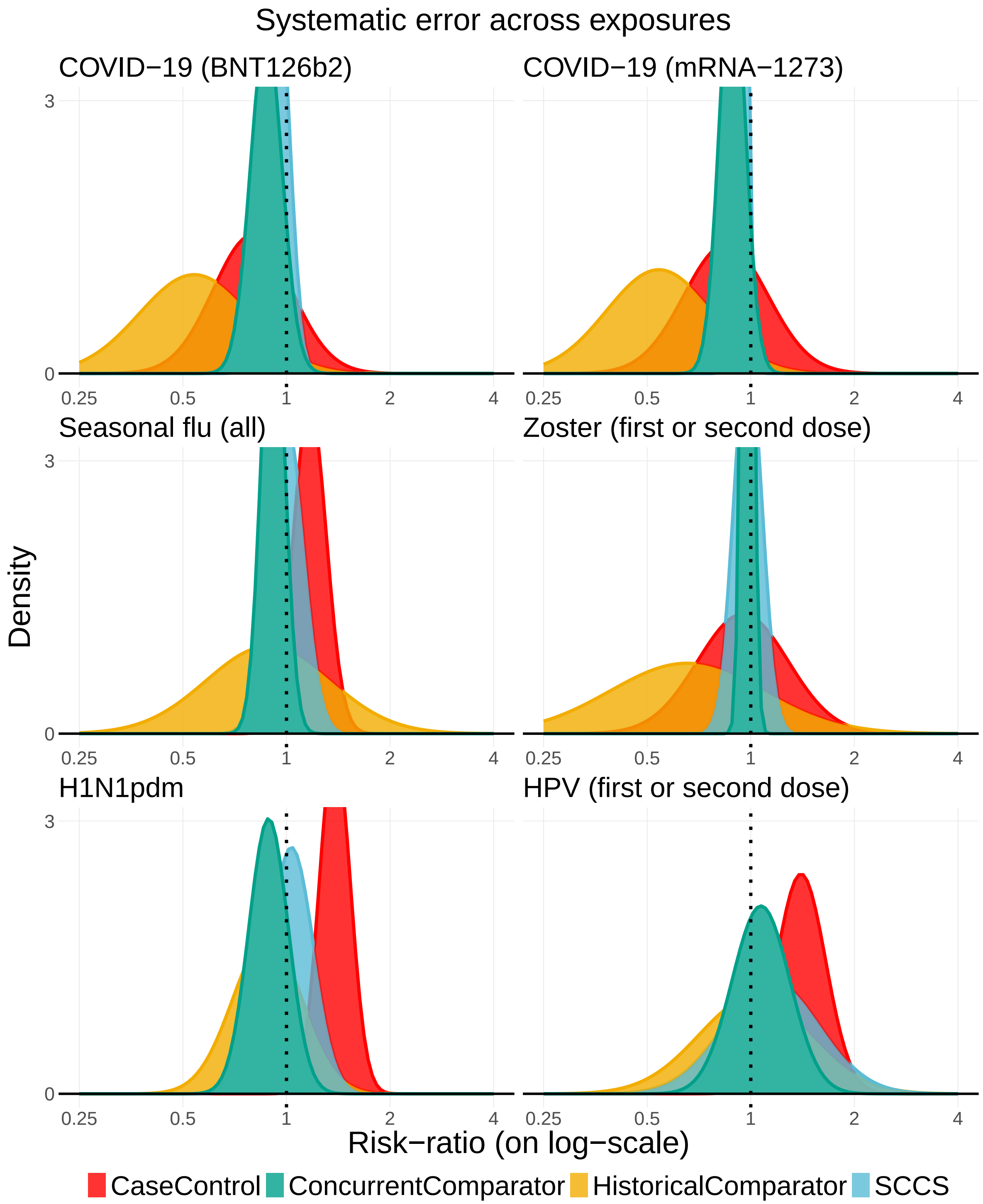


Figure 1: Systematic error distributions of the methods across different exposures for the Optum DOD data source.

#### 1.2 Result for the CCAE data source

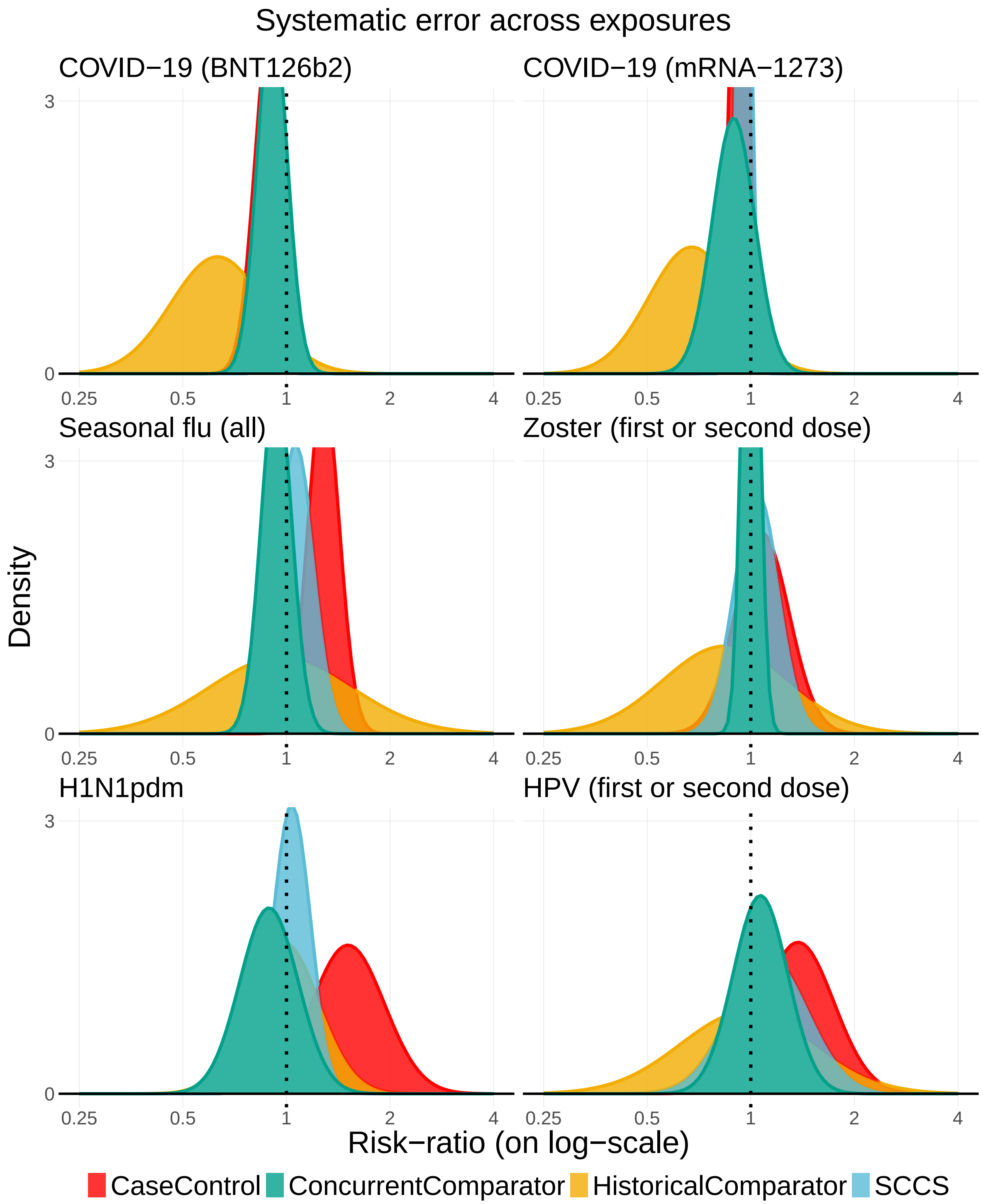


Figure 2: Systematic error distributions of the methods across different exposures for the CCAE data source.

##### 1.3 Results for the MDCD data source

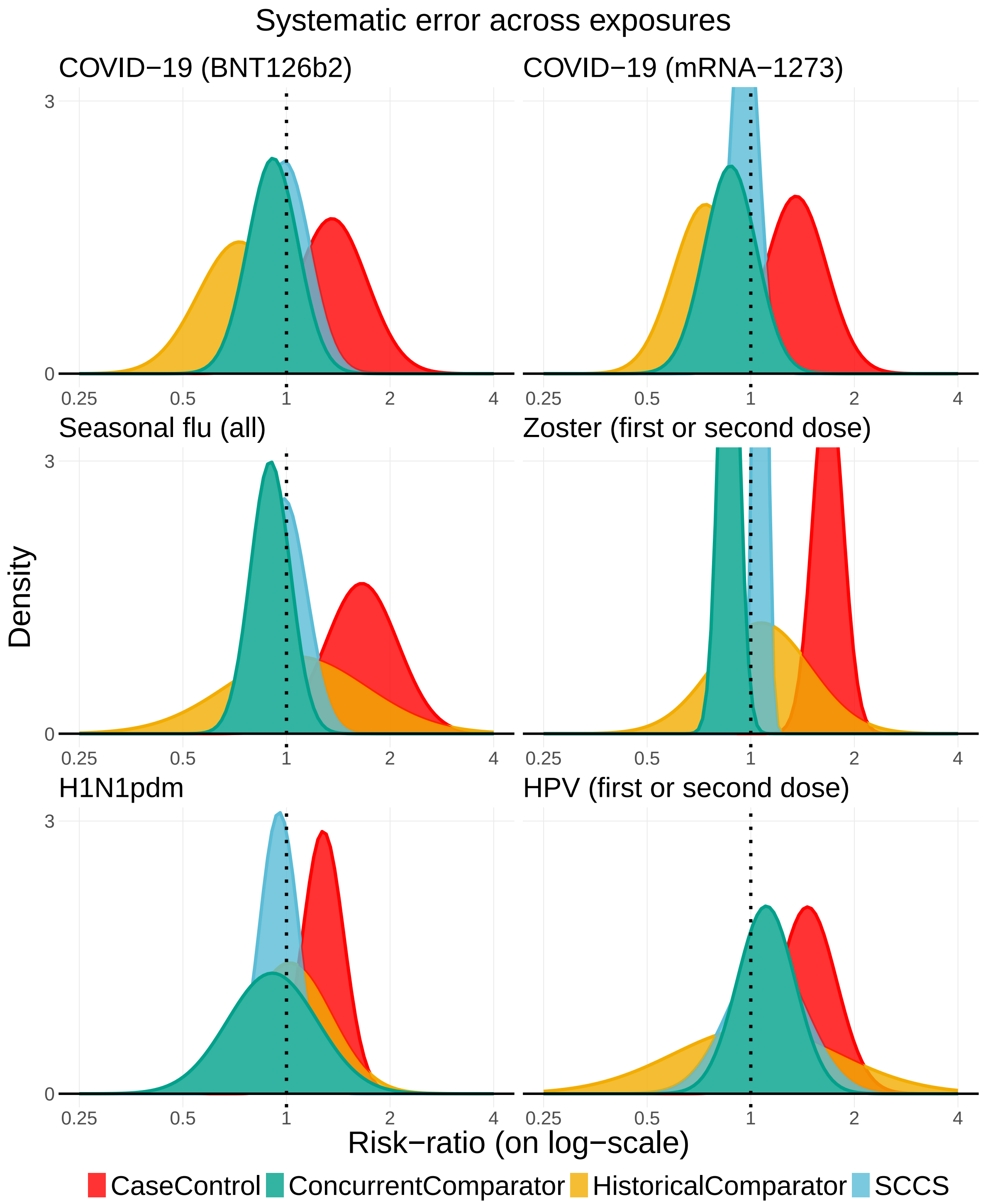


Figure 3: Systematic error distributions of the methods across different exposures for the MDCD data source.

#### 1.4 Results for the MDCR data source

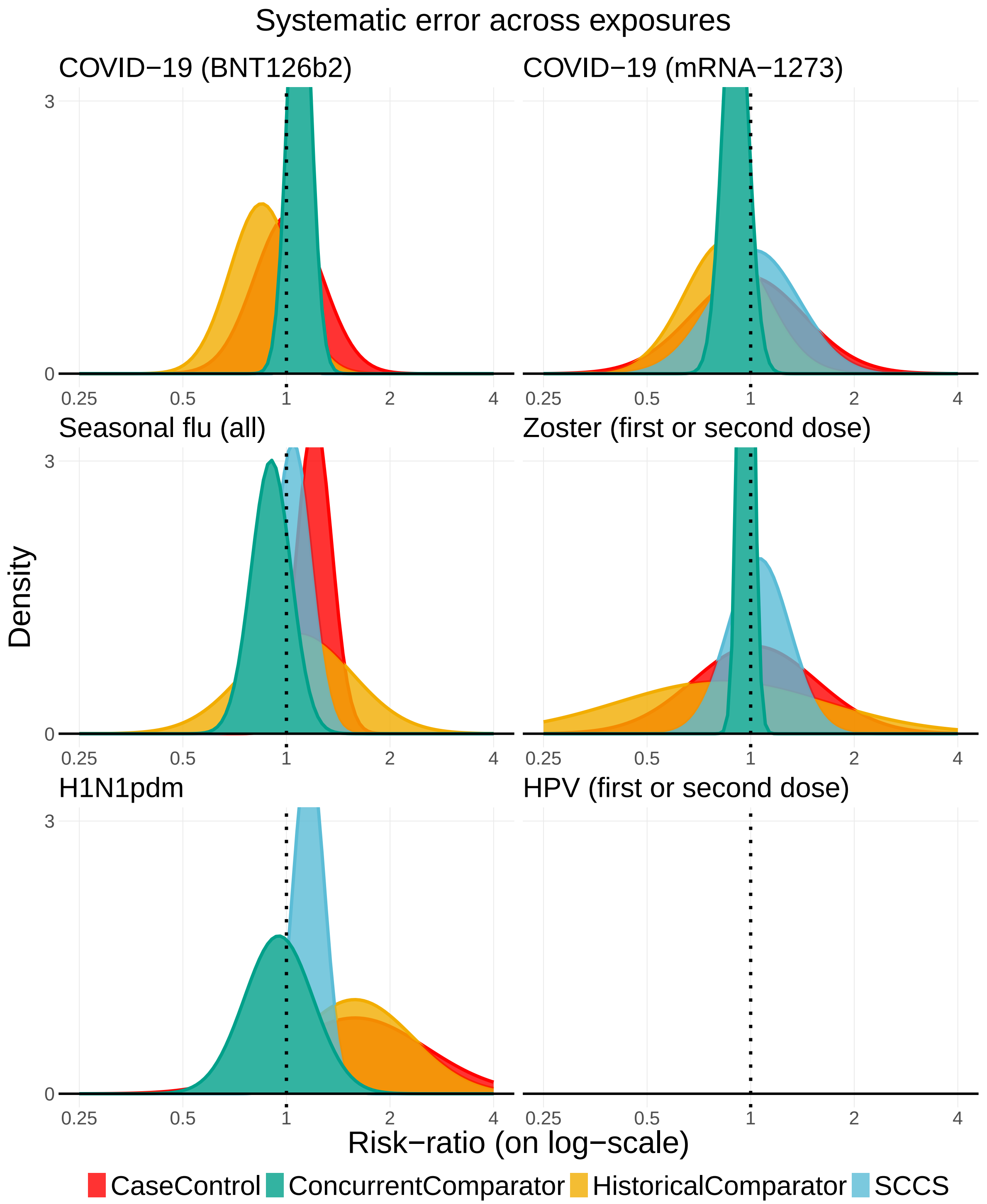


Figure 4: Systematic error distributions of the methods across different exposures for the MDCR data source.

### 2 Type 1 error and power of detection

#### 2.1 Results for Optum EHR data source

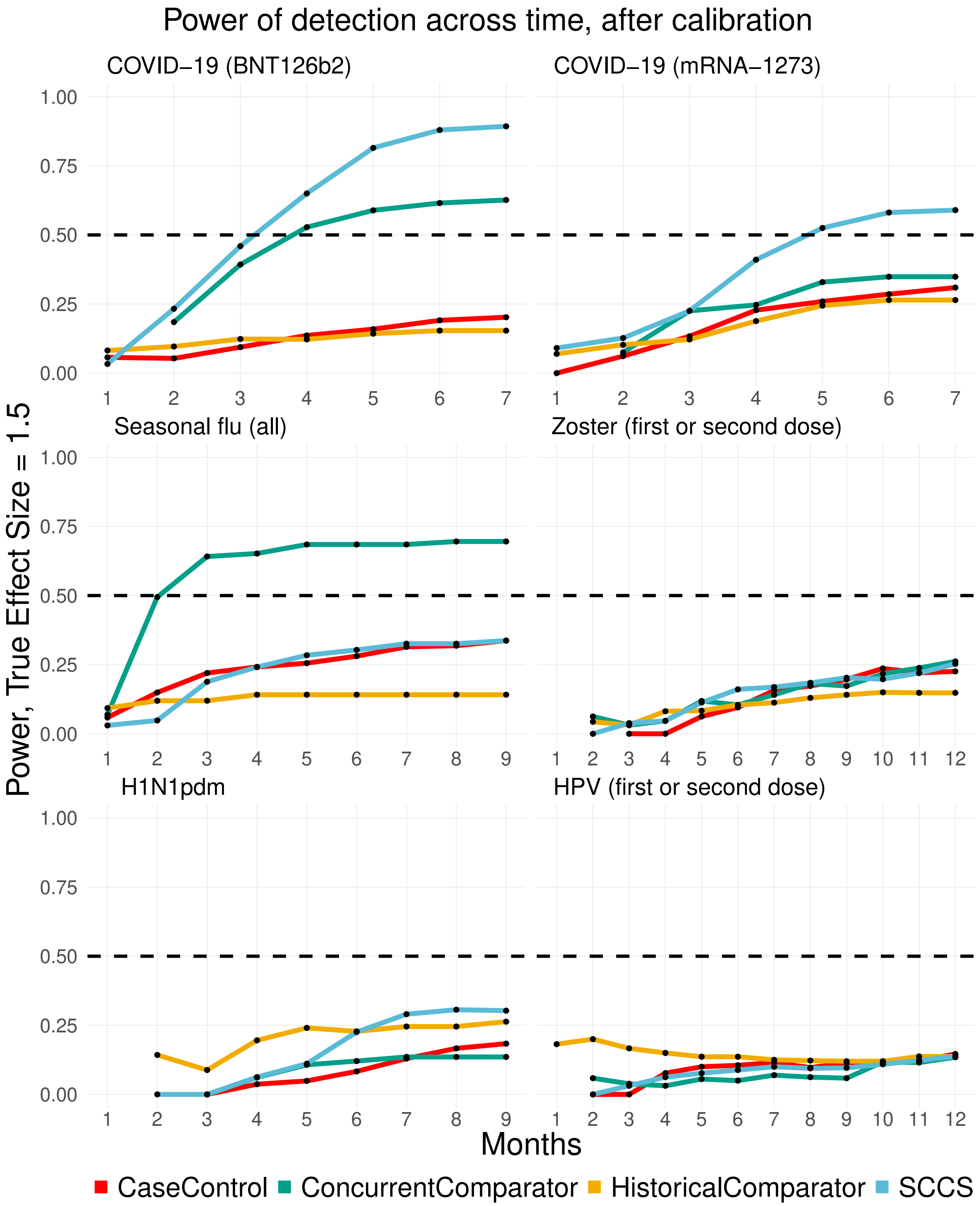


Figure 5: Power of detection (true effect size = 1.5) of the methods as a function of time, across different exposures for the Optum EHR data source.

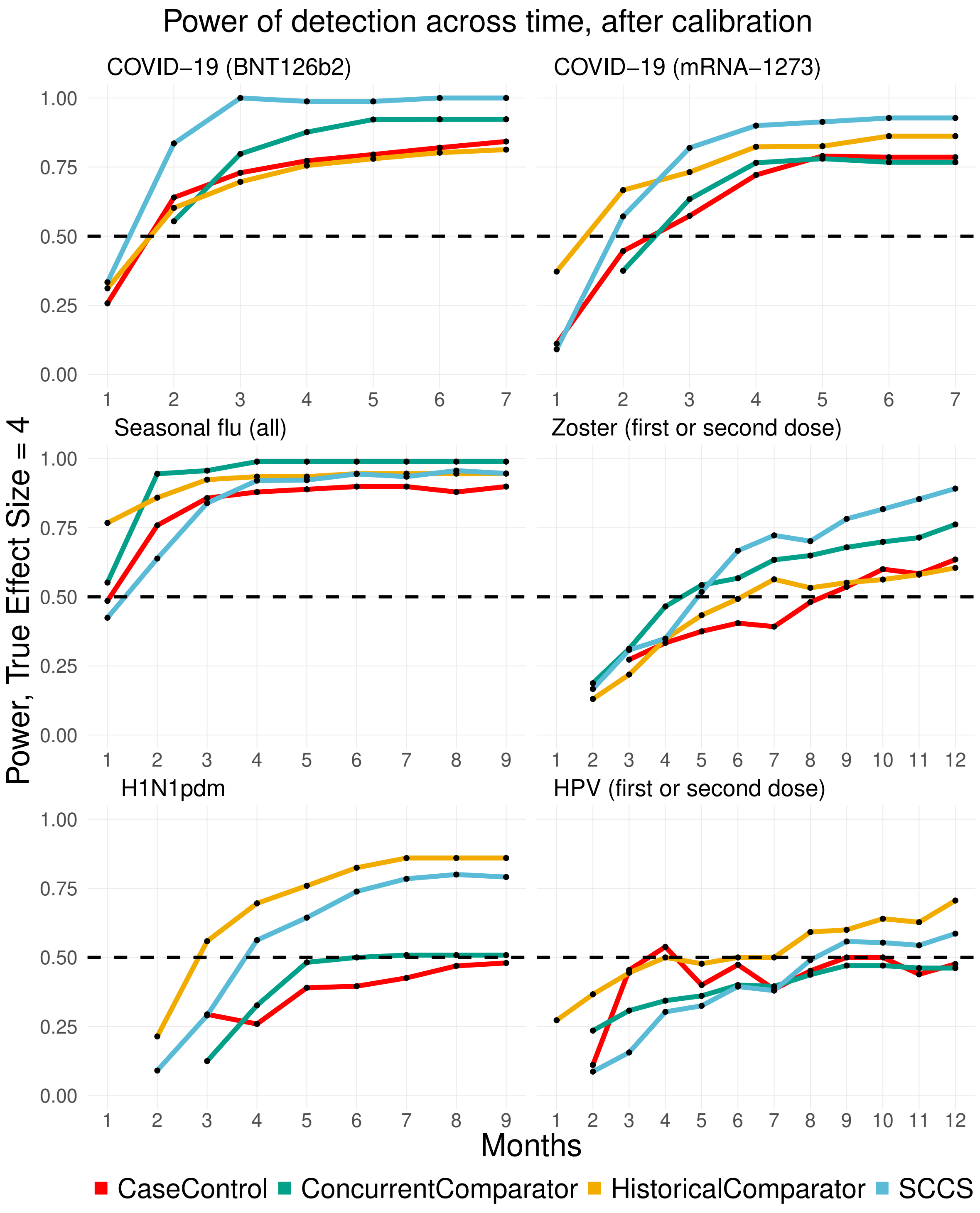


Figure 6: Power of detection (true effect size = 4) of the methods as a function of time, across different exposures for the Optum EHR data source.

#### 2.2 Results for Optum DOD data source

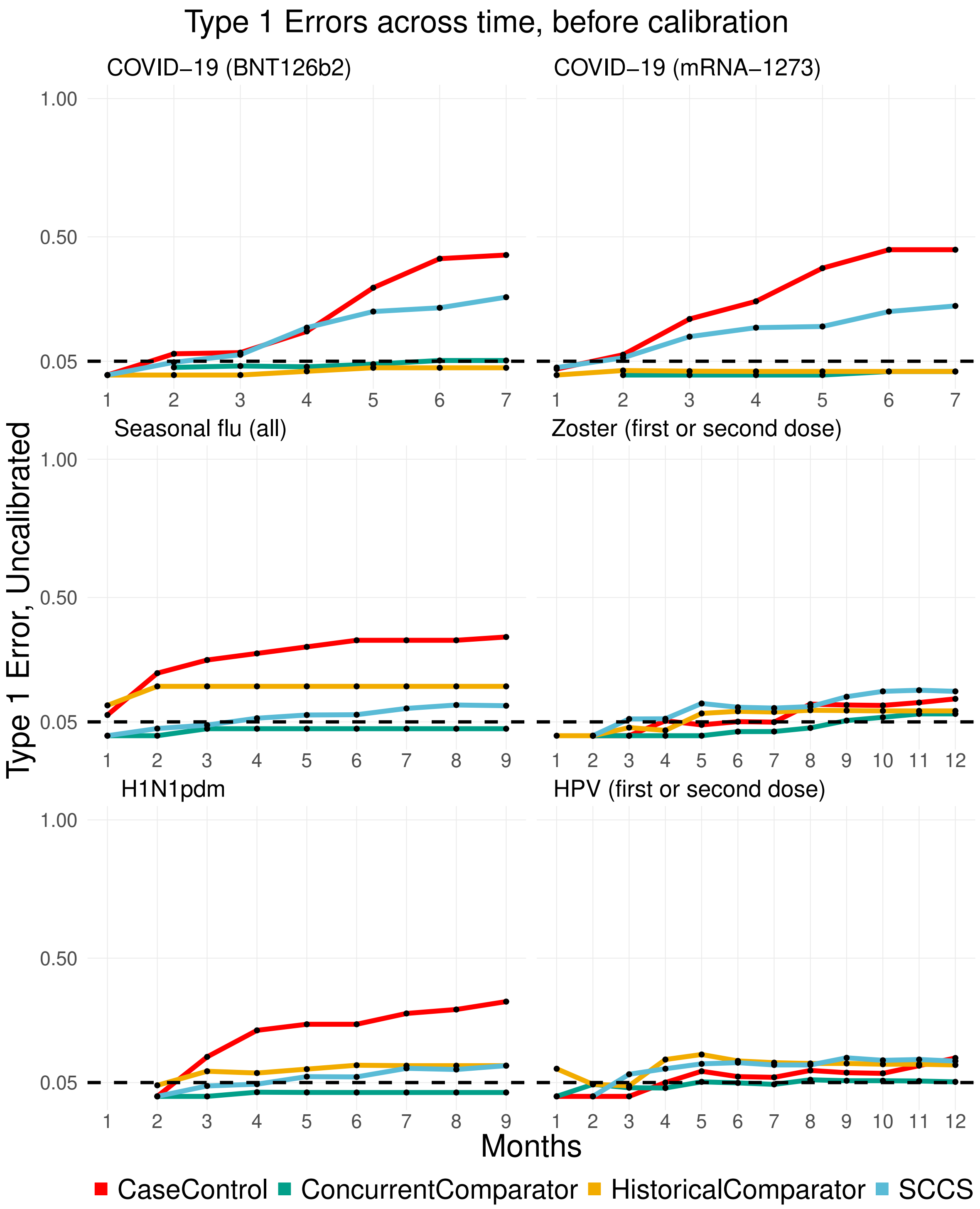


Figure 7: Uncalibrated Type 1 errors of methods as a function of time, across different exposures for the Optum DOD data source.

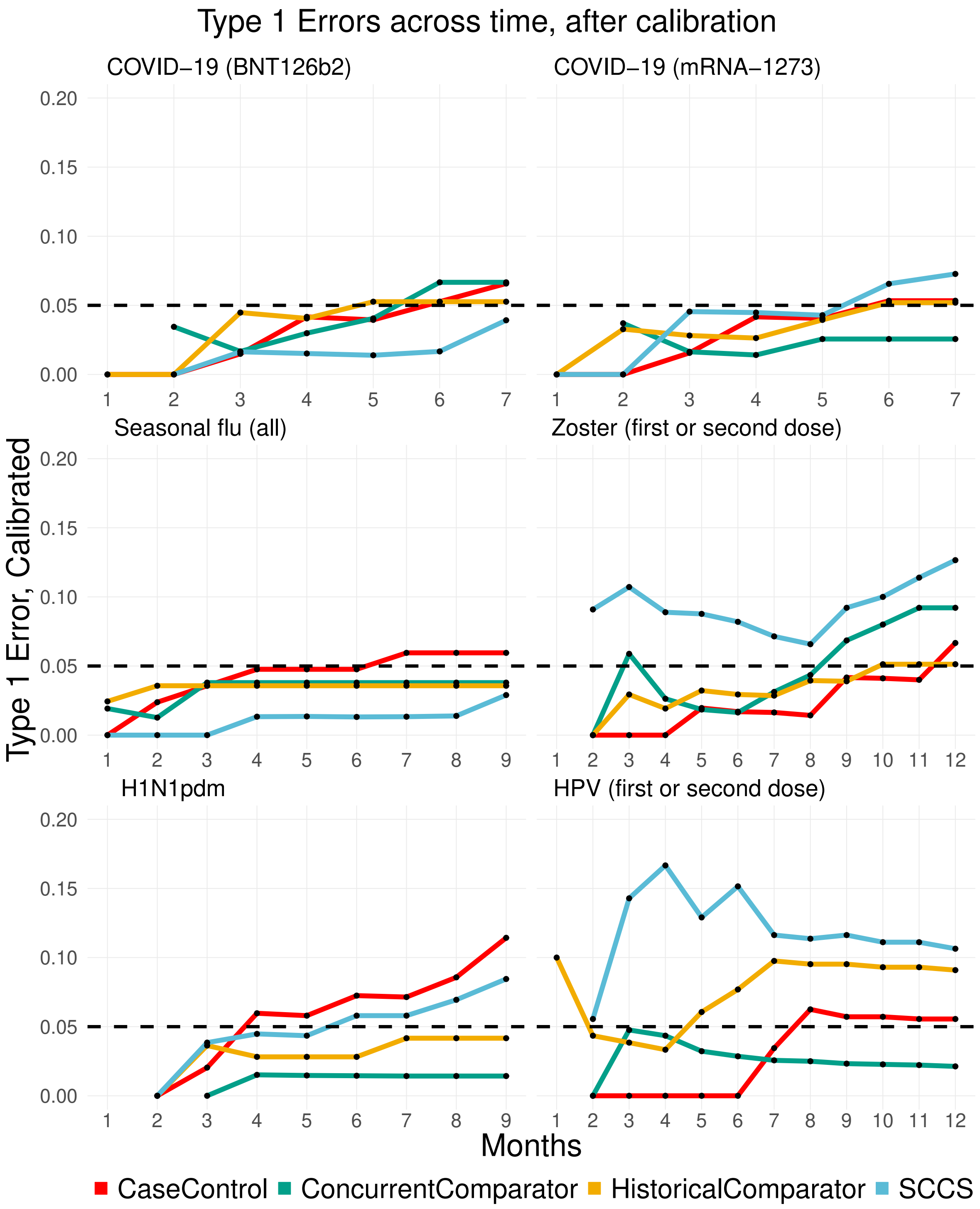


Figure 8: Calibrated Type 1 errors of methods as a function of time, across different exposures for the Optum DOD data source.

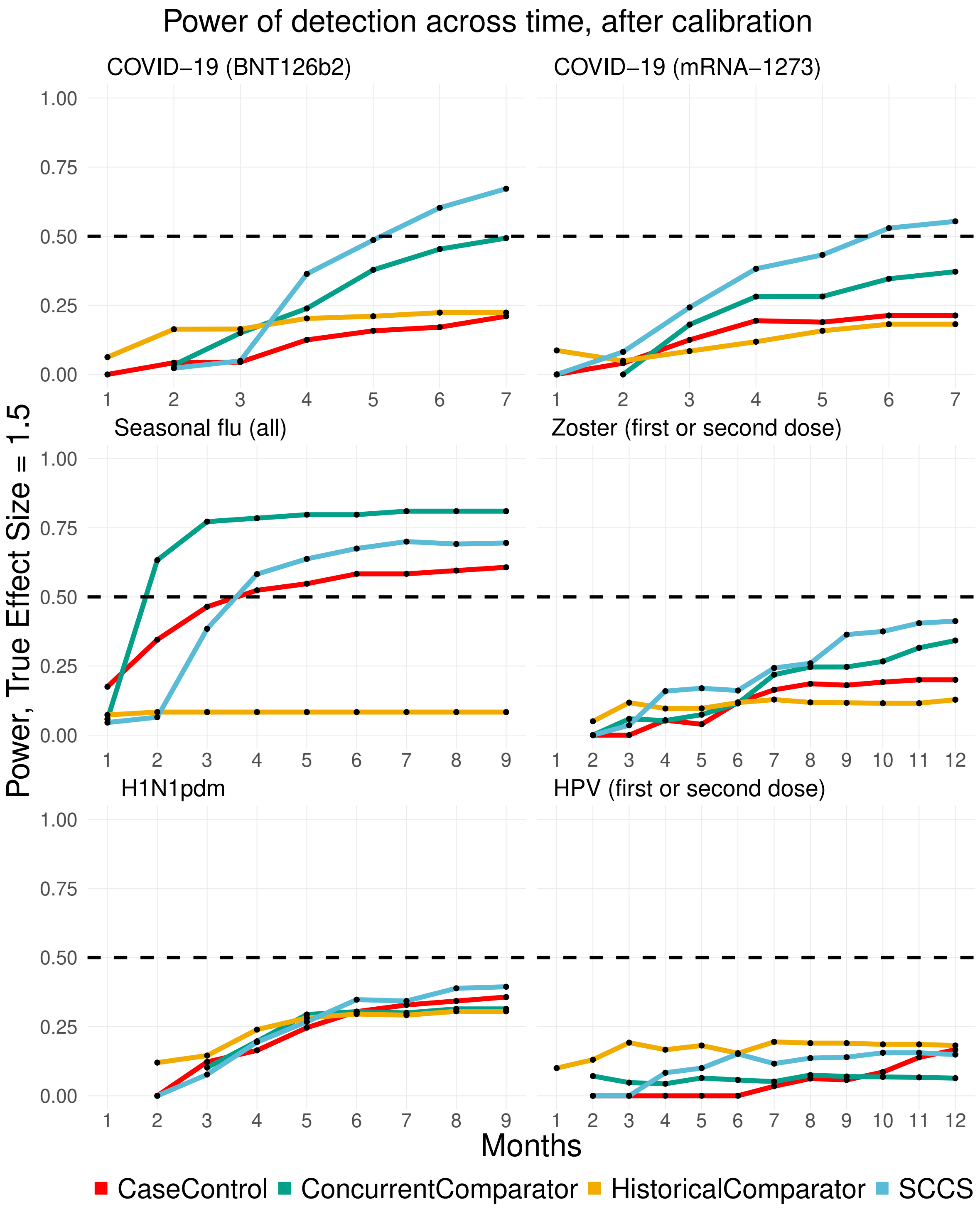


Figure 9: Power of detection (true effect size = 1.5) of the methods as a function of time, across different exposures for the Optum DOD data source.

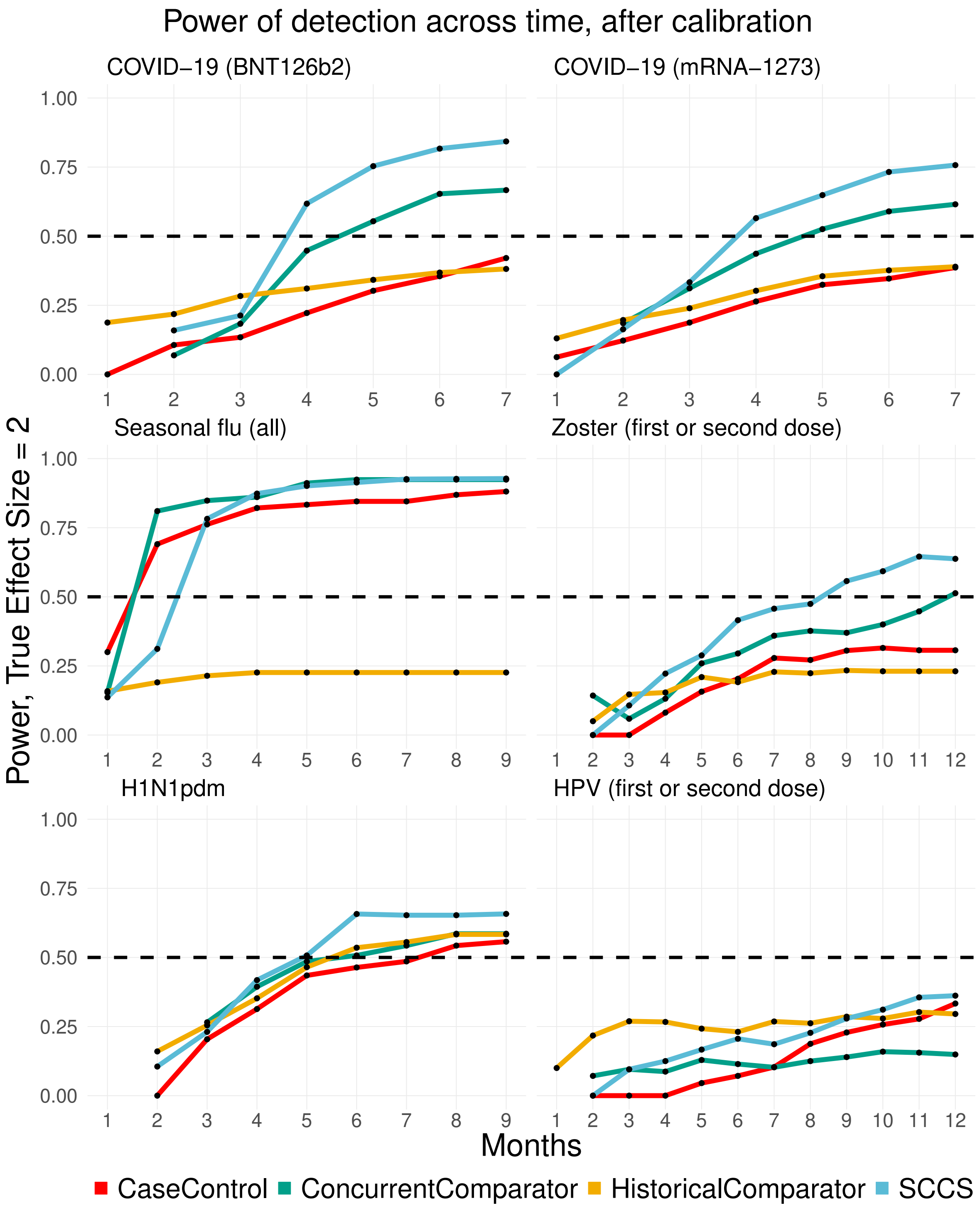


Figure 10: Power of detection (true effect size = 2) of the methods as a function of time, across different exposures for the Optum DOD data source.

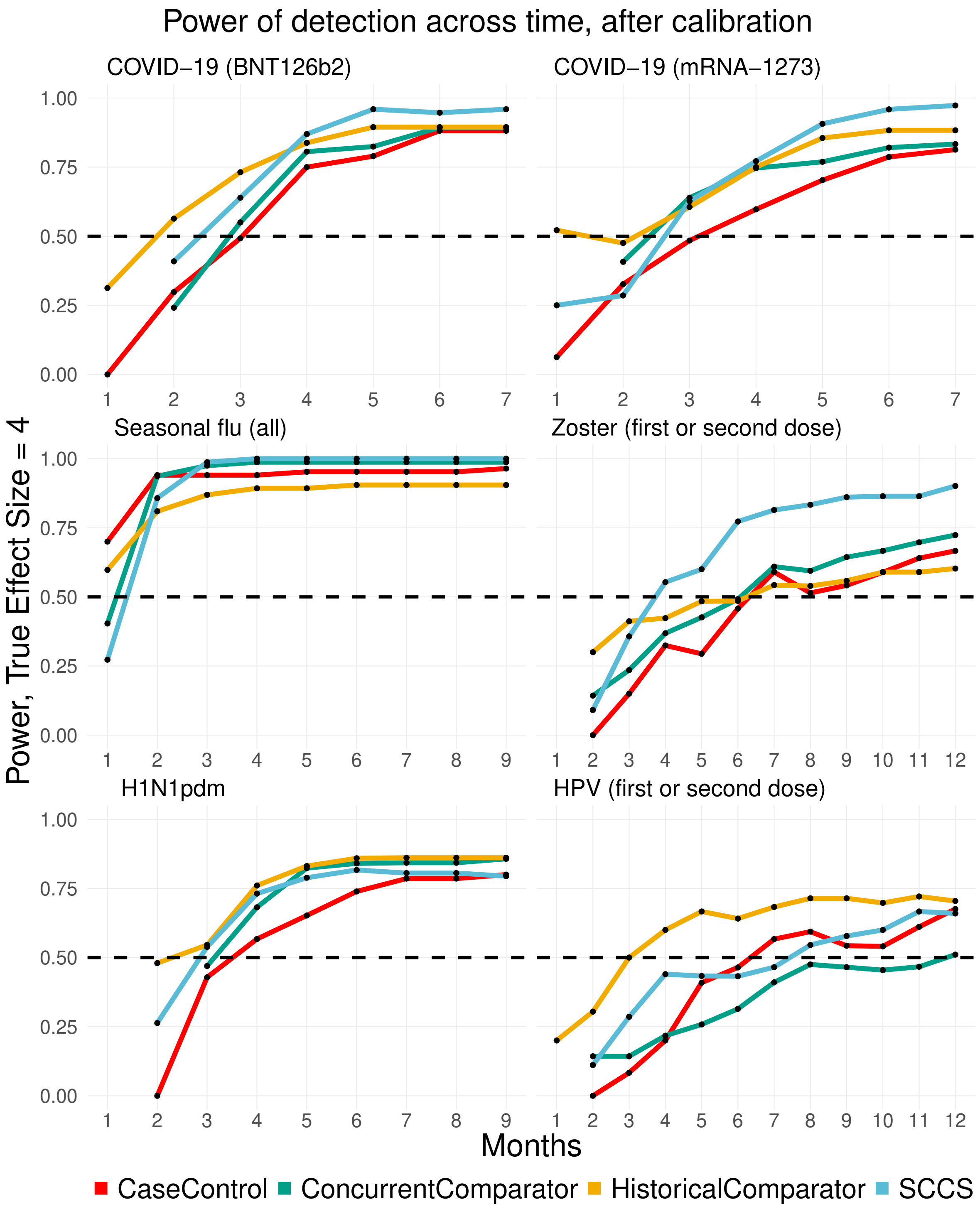


Figure 11: Power of detection (true effect size = 4) of the methods as a function of time, across different exposures for the Optum DOD data source.

#### 2.3 Results for CCAE data source

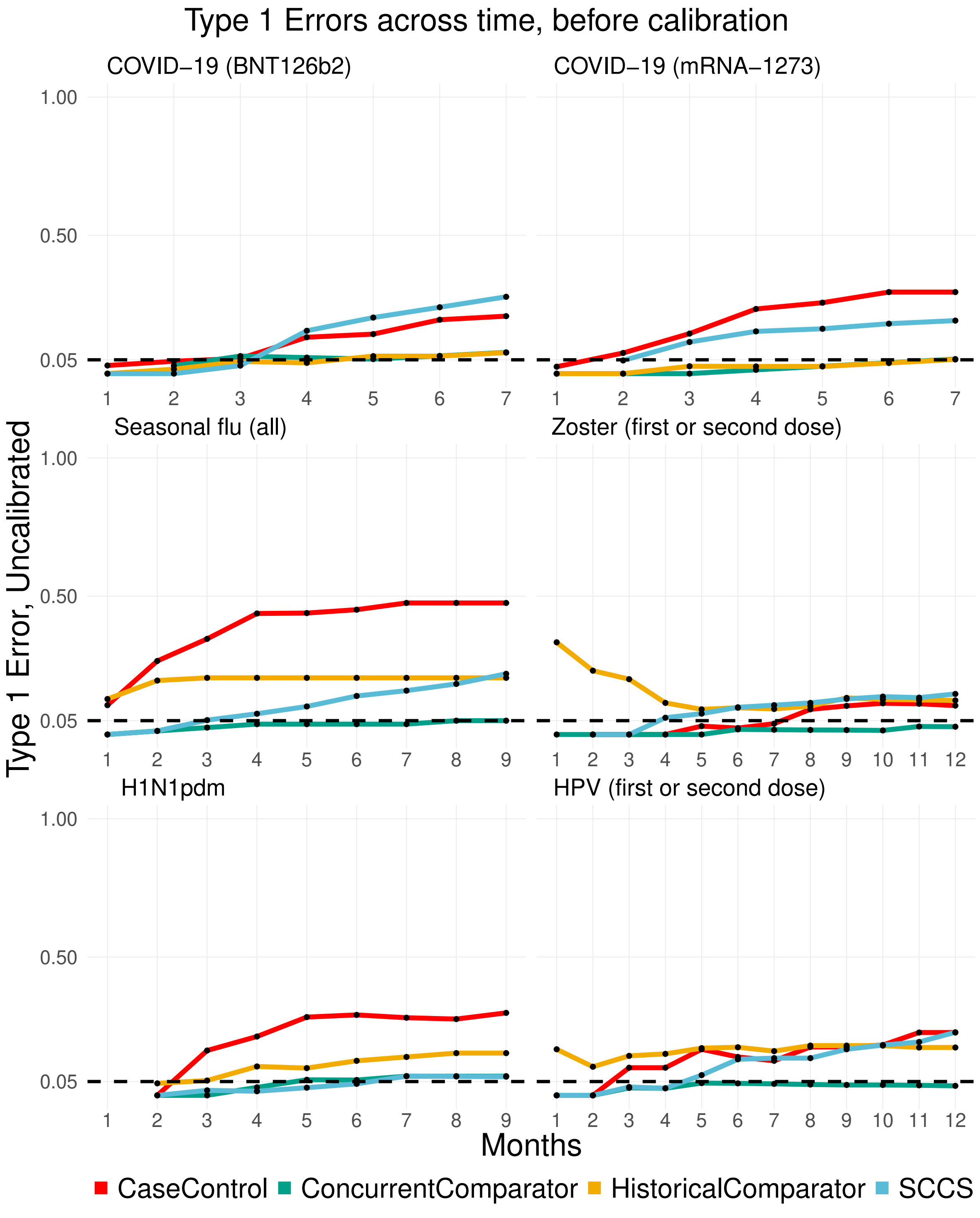


Figure 12: Uncalibrated Type 1 errors of methods as a function of time, across different exposures for the CCAE data source.

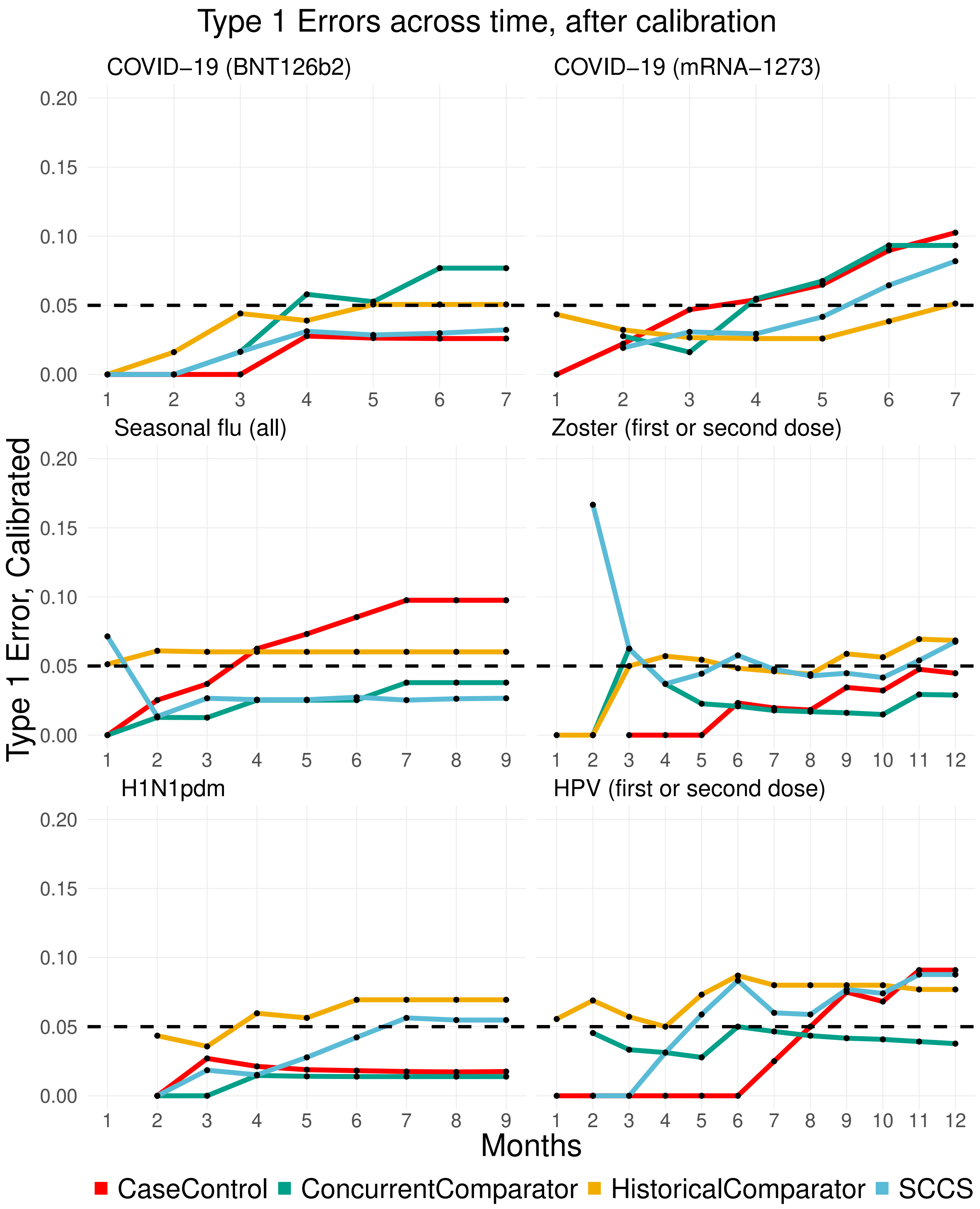


Figure 13: Calibrated Type 1 errors of methods as a function of time, across different exposures for the CCAE data source.

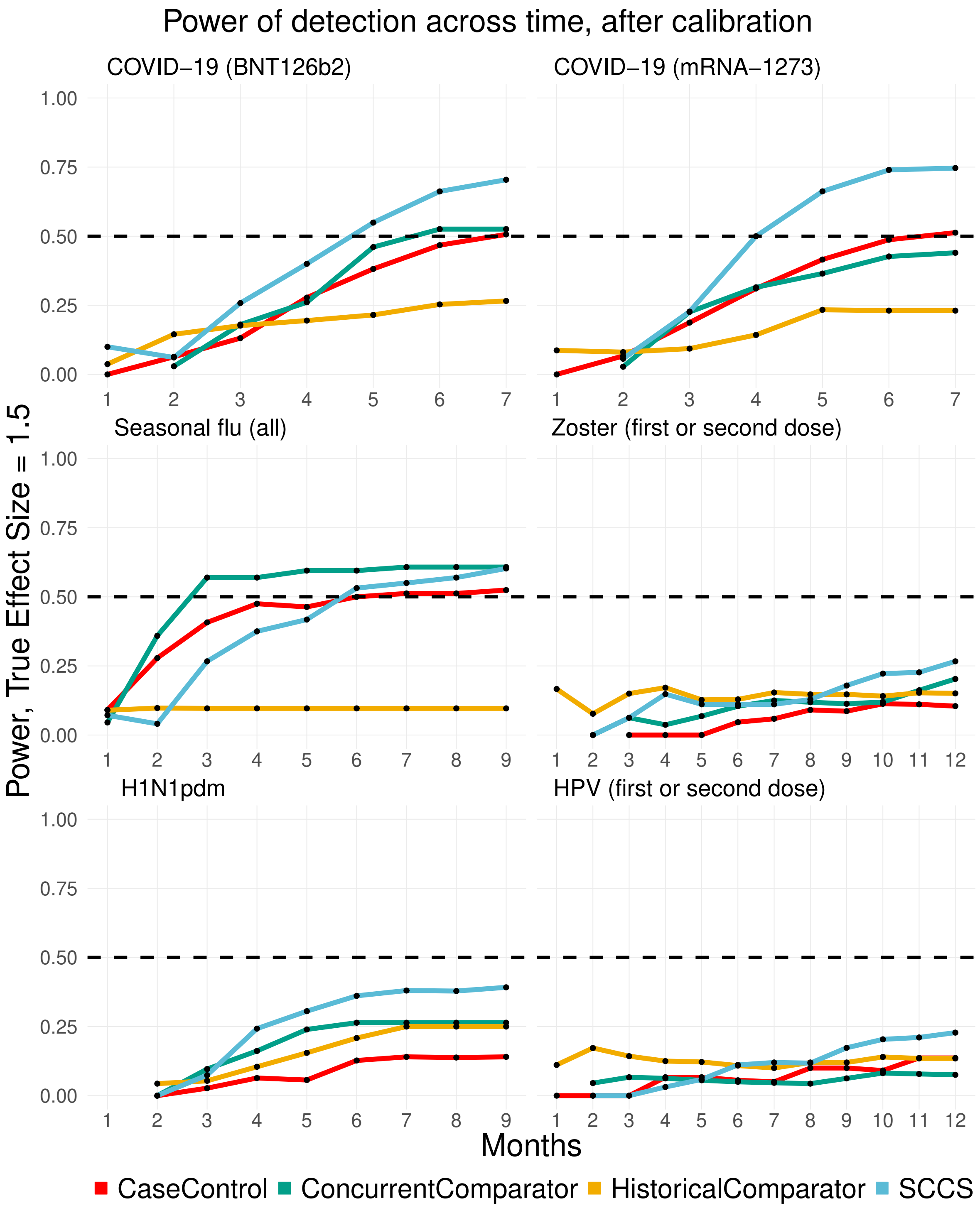


Figure 14: Power of detection (true effect size = 1.5) of the methods as a function of time, across different exposures for the CCAE data source.

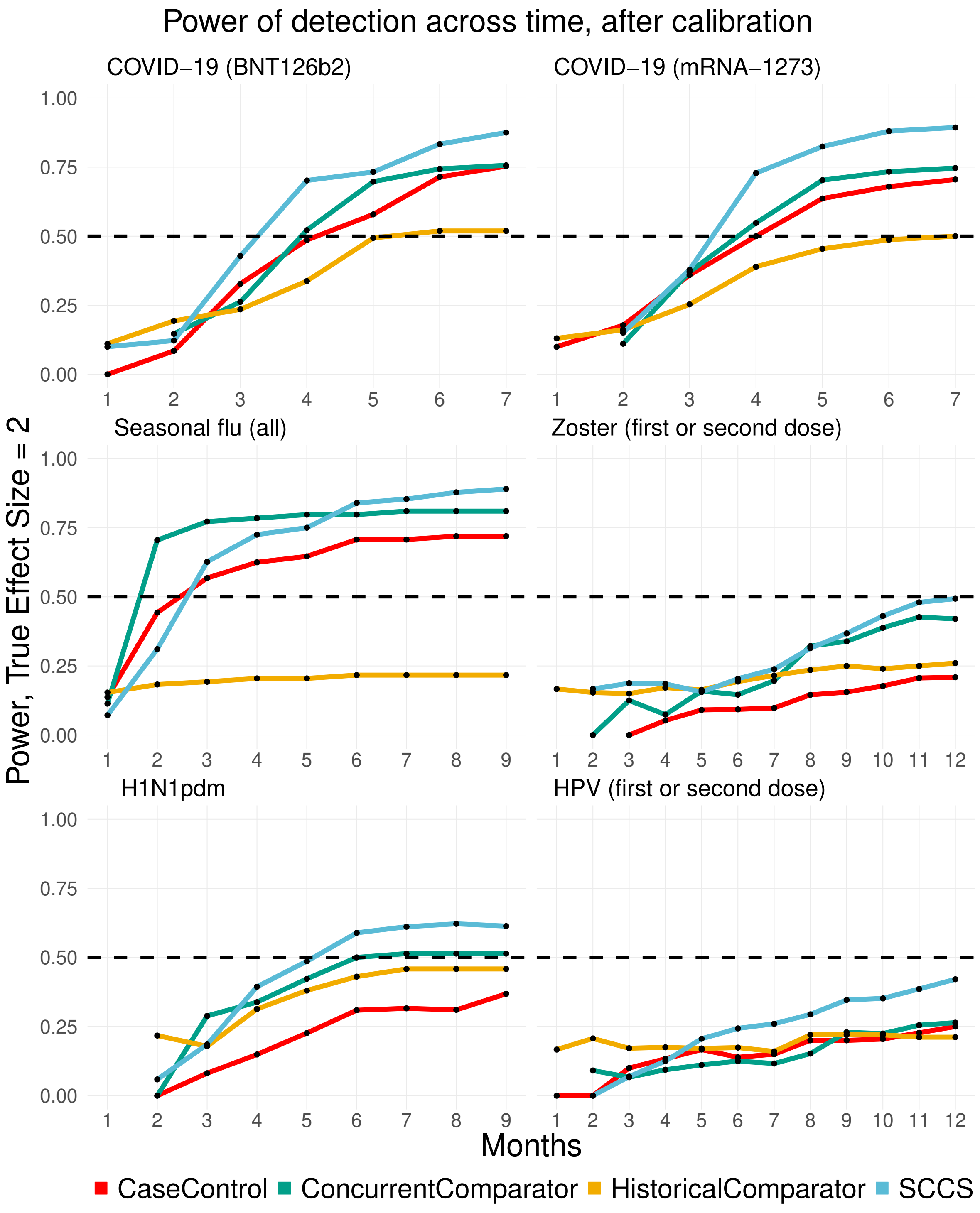


Figure 15: Power of detection (true effect size = 2) of the methods as a function of time, across different exposures for the CCAE data source.

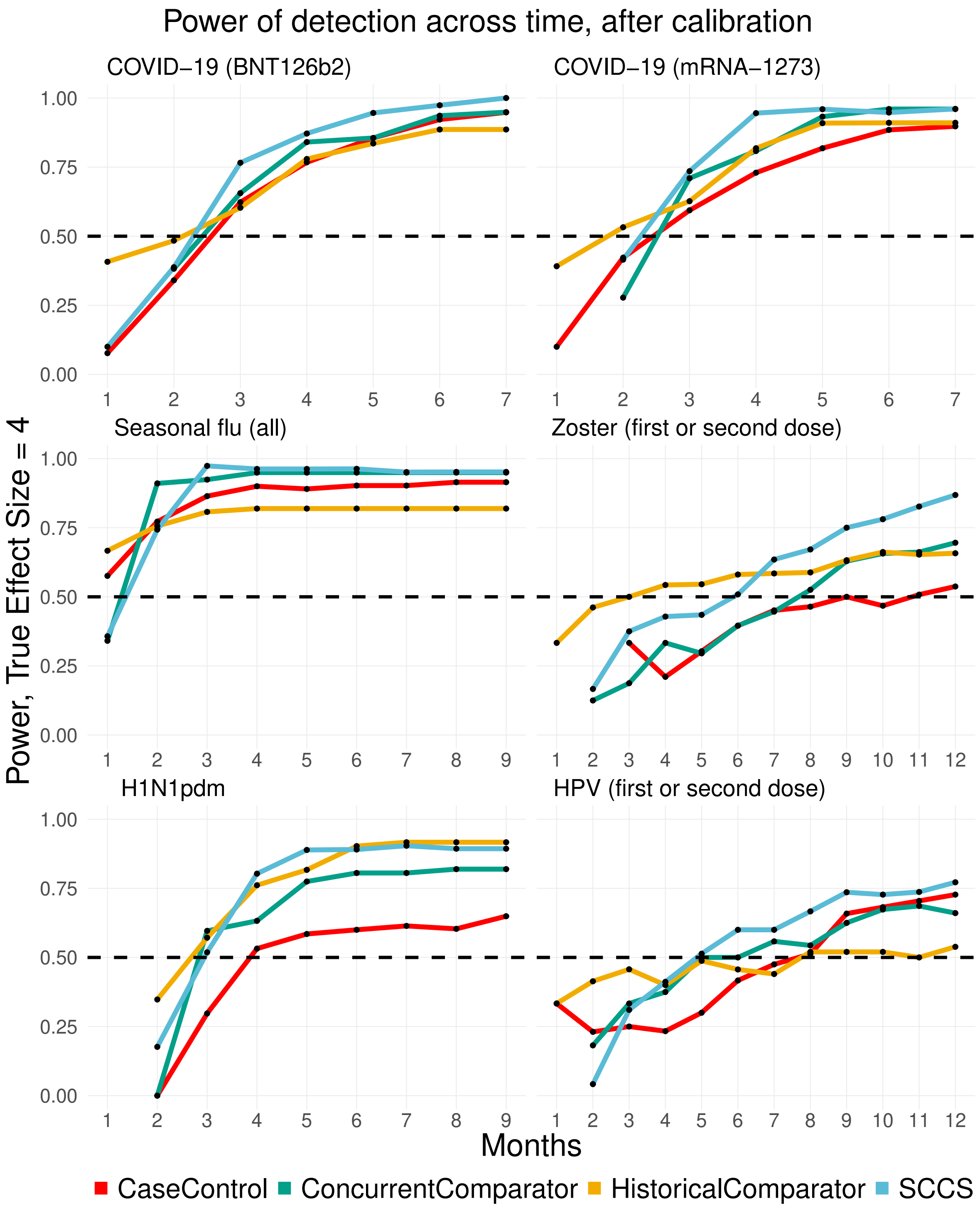


Figure 16: Power of detection (true effect size = 4) of the methods as a function of time, across different exposures for the CCAE data source.

#### 2.4 Results for MDCD data source

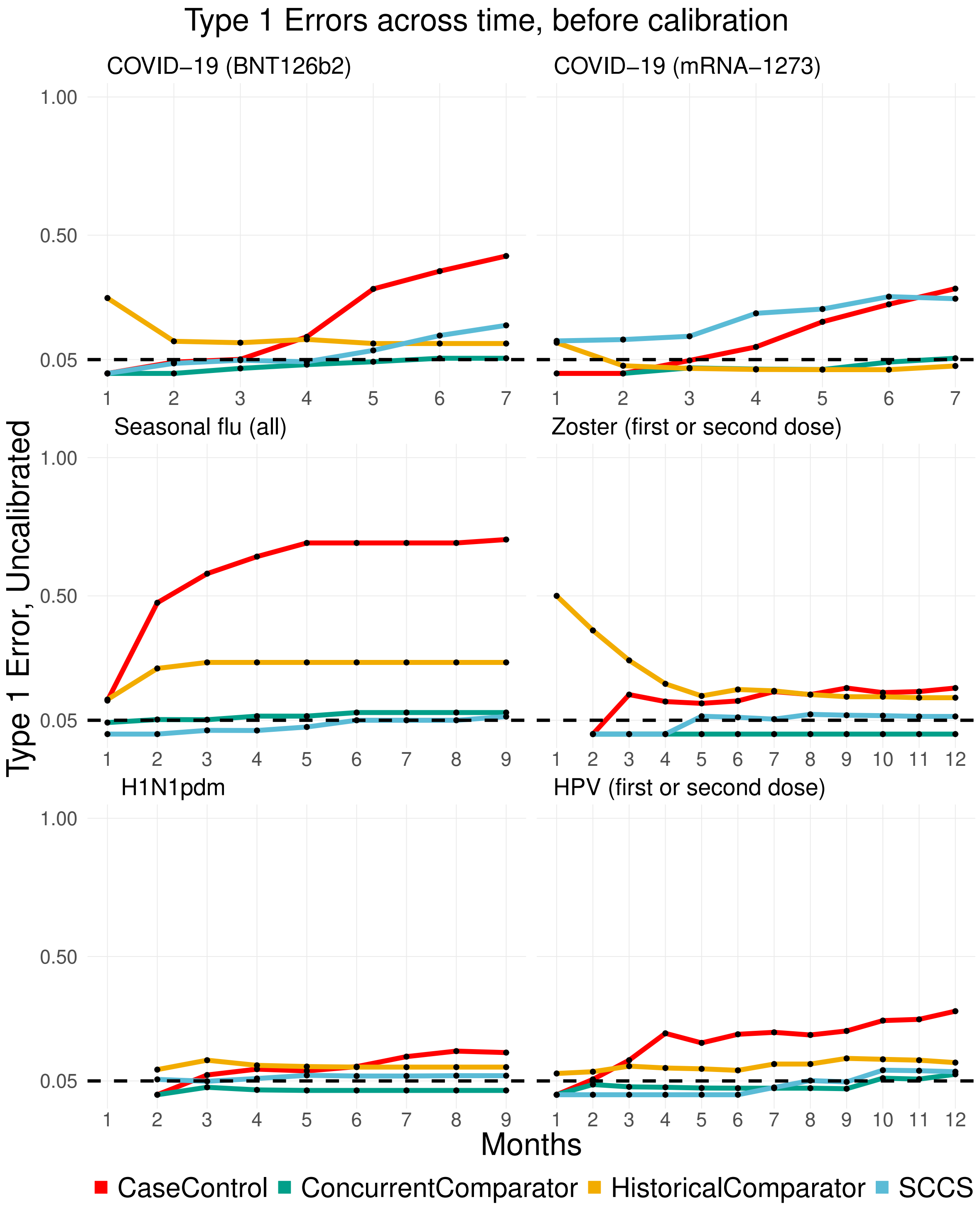


Figure 17: Uncalibrated Type 1 errors of methods as a function of time, across different exposures for the MDCD data source.

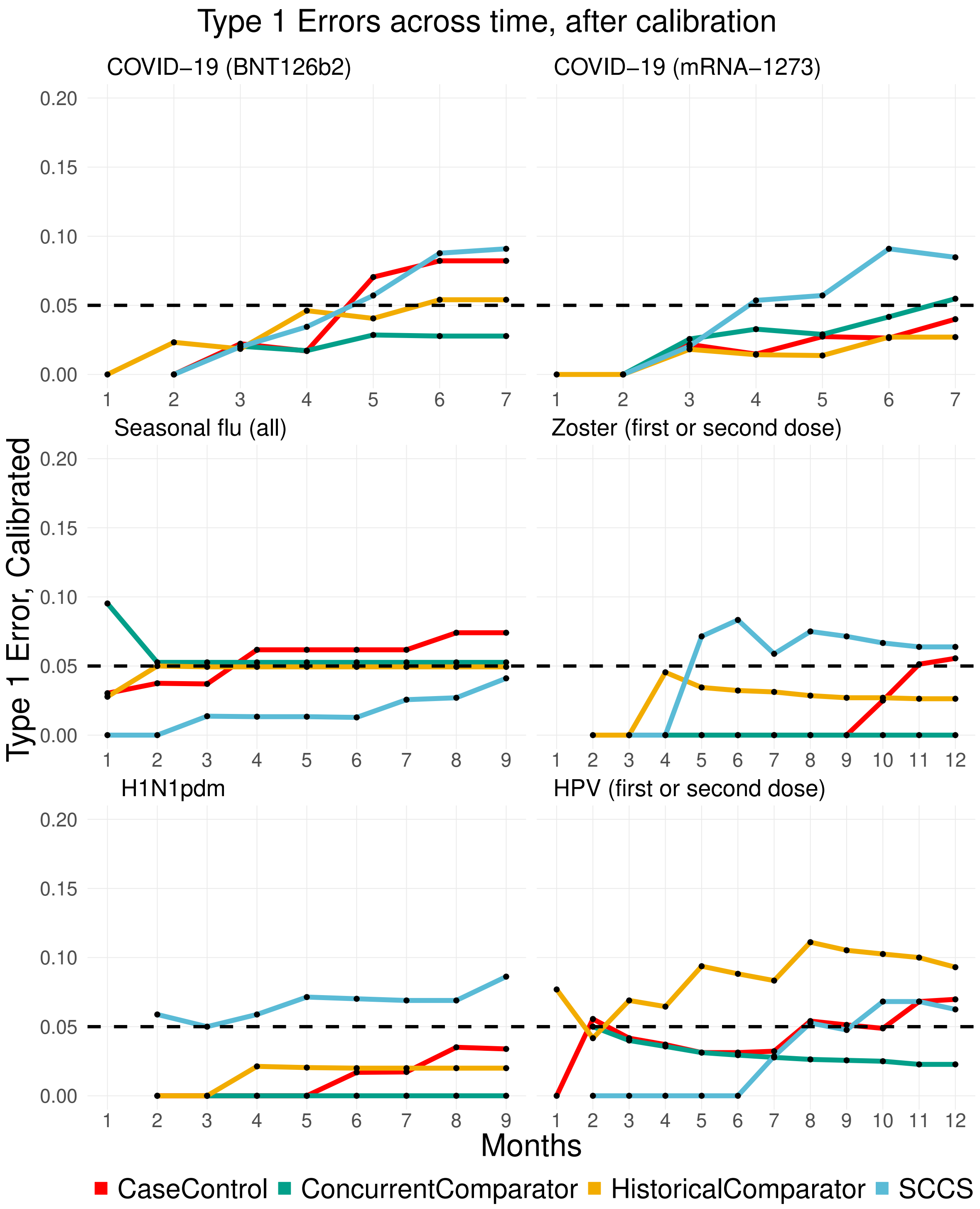


Figure 18: Calibrated Type 1 errors of methods as a function of time, across different exposures for the MDCD data source.

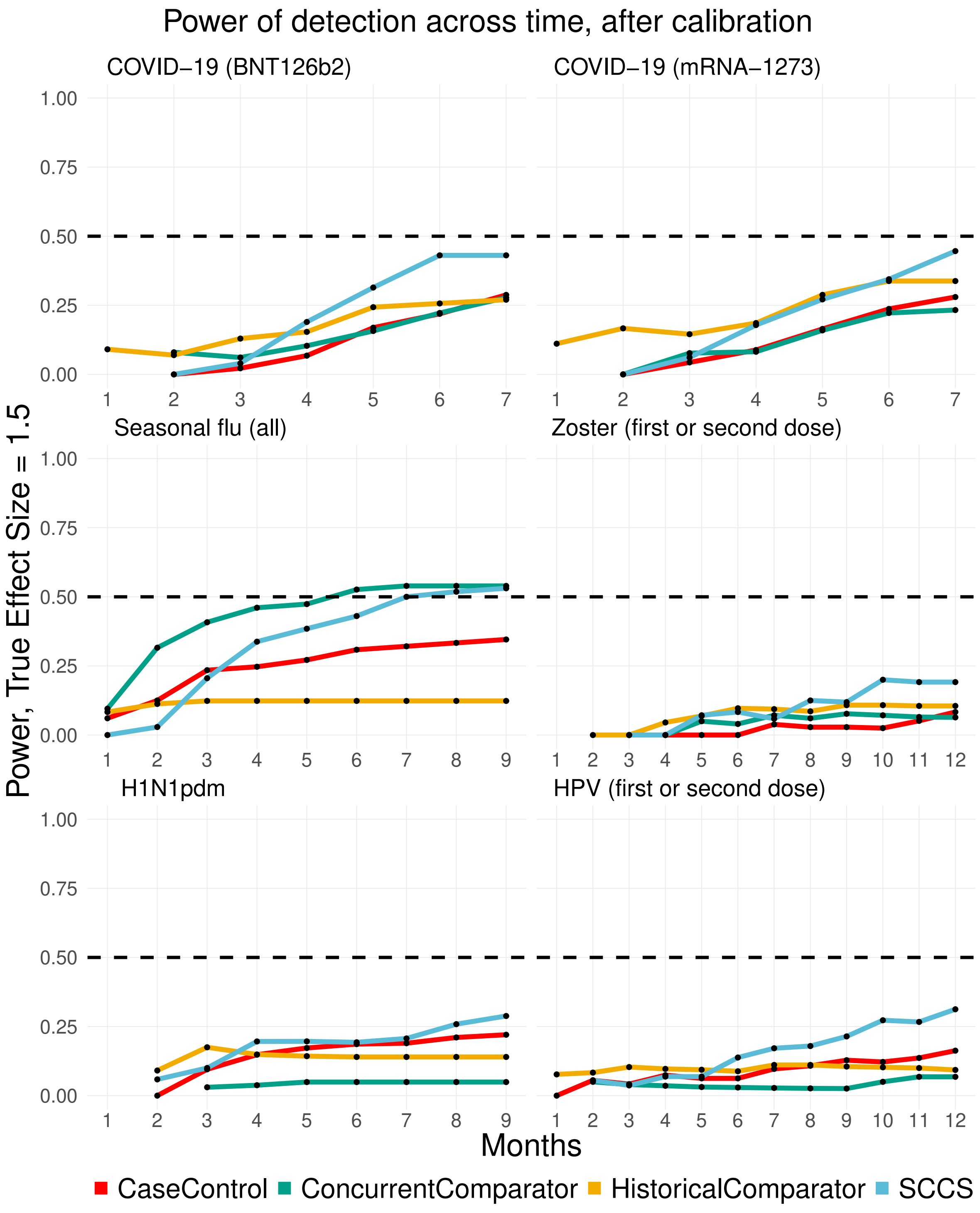


Figure 19: Power of detection (true effect size = 1.5) of the methods as a function of time, across different exposures for the MDCD data source.

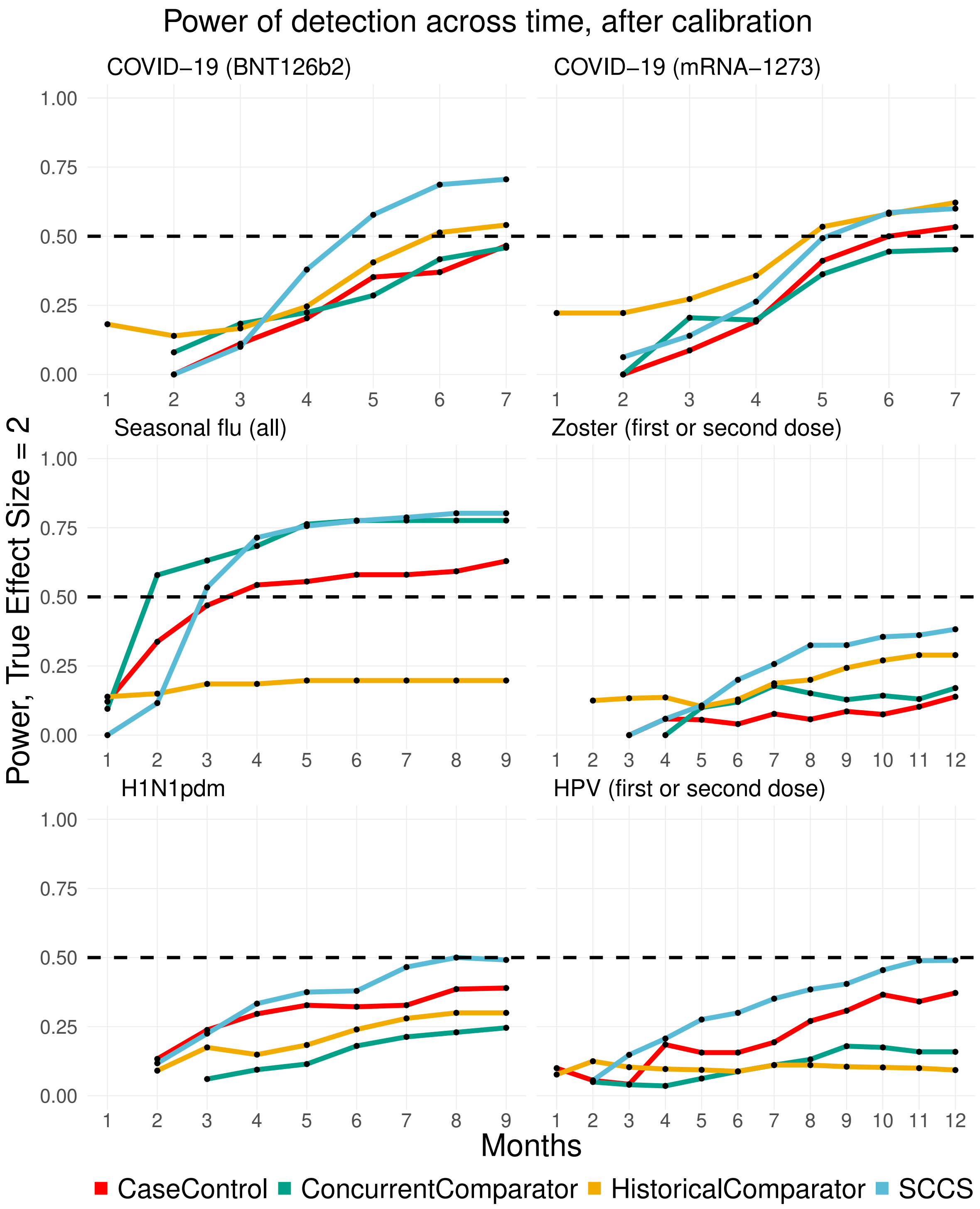


Figure 20: Power of detection (true effect size = 2) of the methods as a function of time, across different exposures for the MDCD data source.

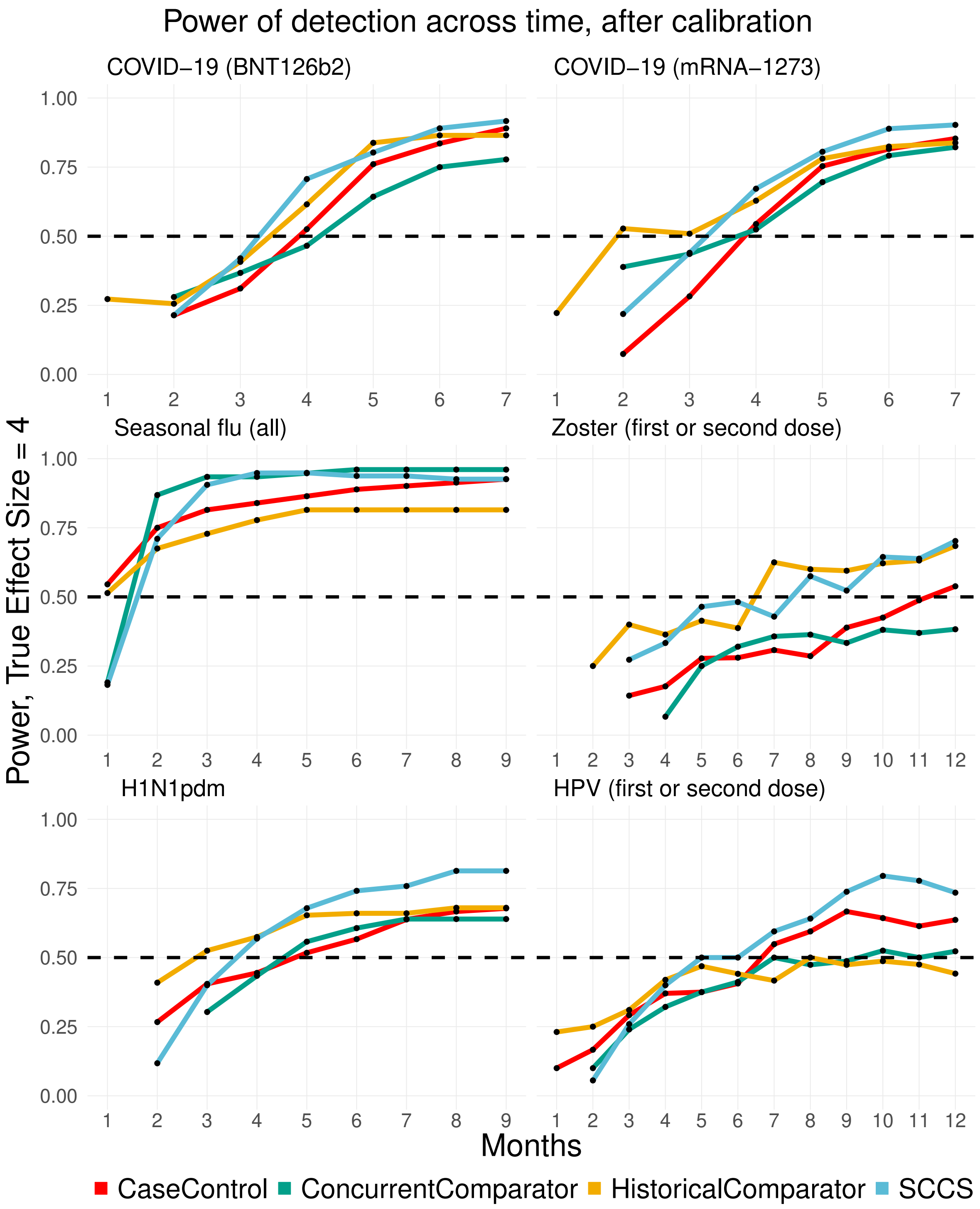


Figure 21: Power of detection (true effect size = 4) of the methods as a function of time, across different exposures for the MDCD data source.

##### 2.5 Results for MDCR data source

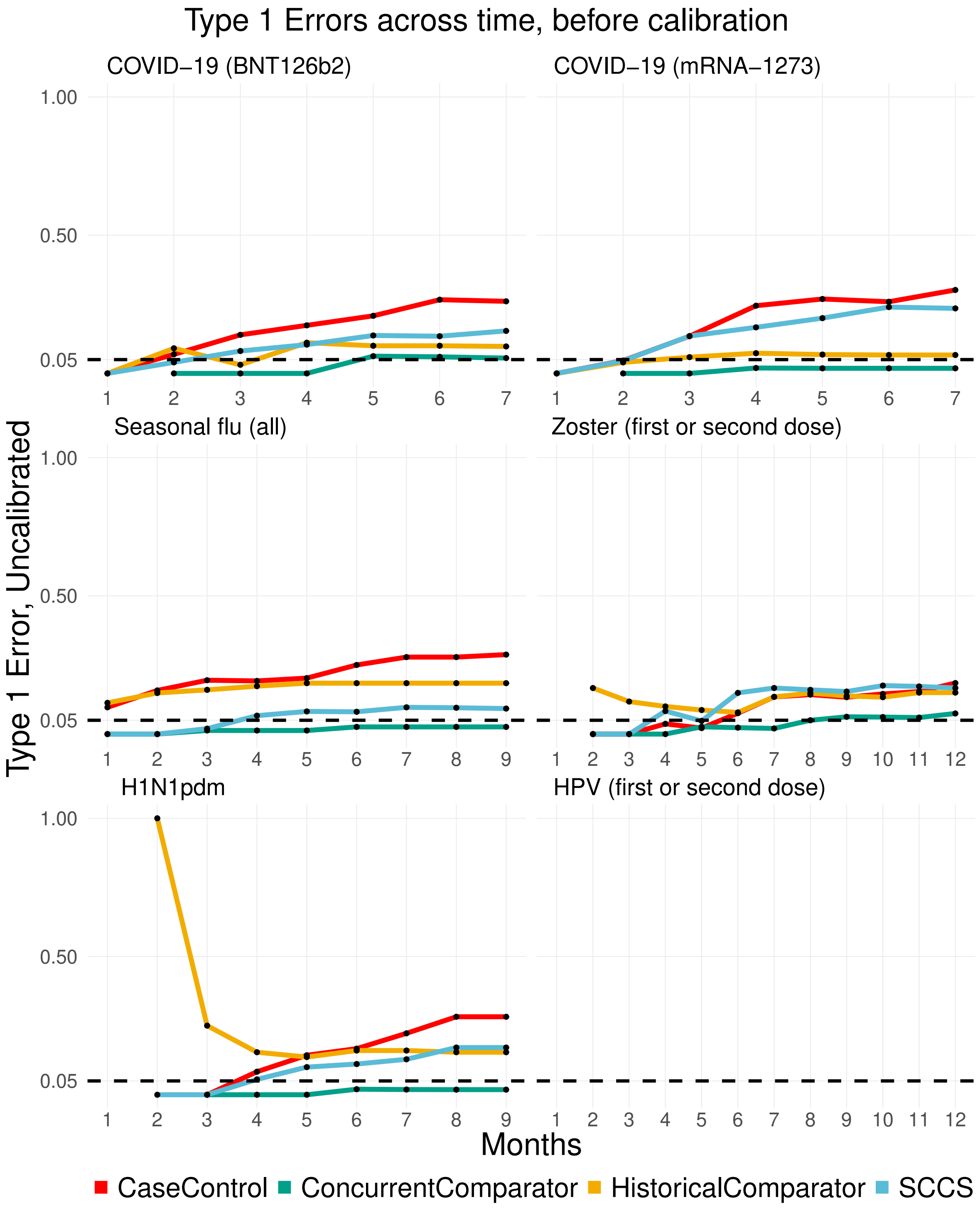


Figure 22: Uncalibrated Type 1 errors of methods as a function of time, across different exposures for the MDCR data source.

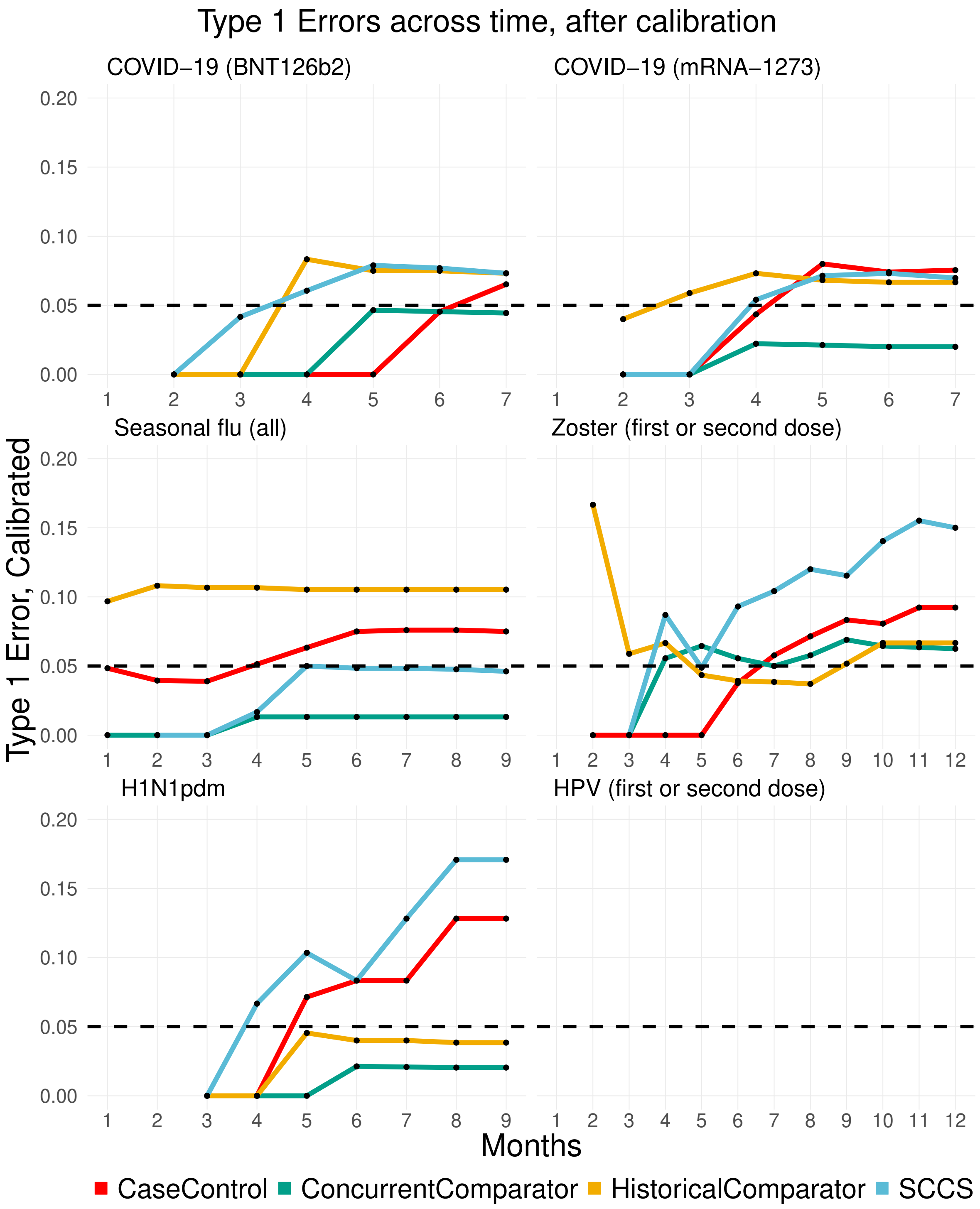


Figure 23: Calibrated Type 1 errors of methods as a function of time, across different exposures for the MDCR data source.

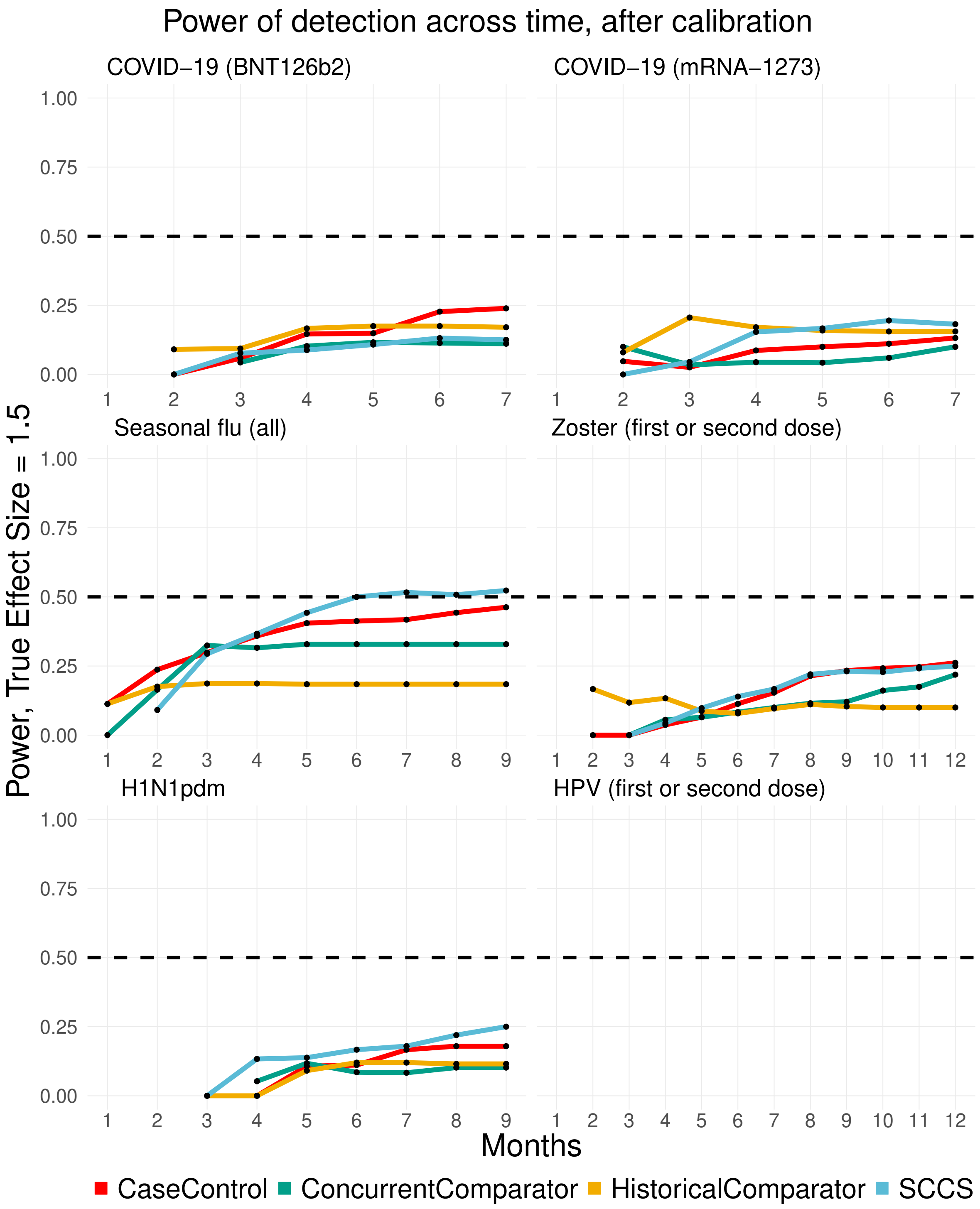


Figure 24: Power of detection (true effect size = 1.5) of the methods as a function of time, across different exposures for the MDCR data source.

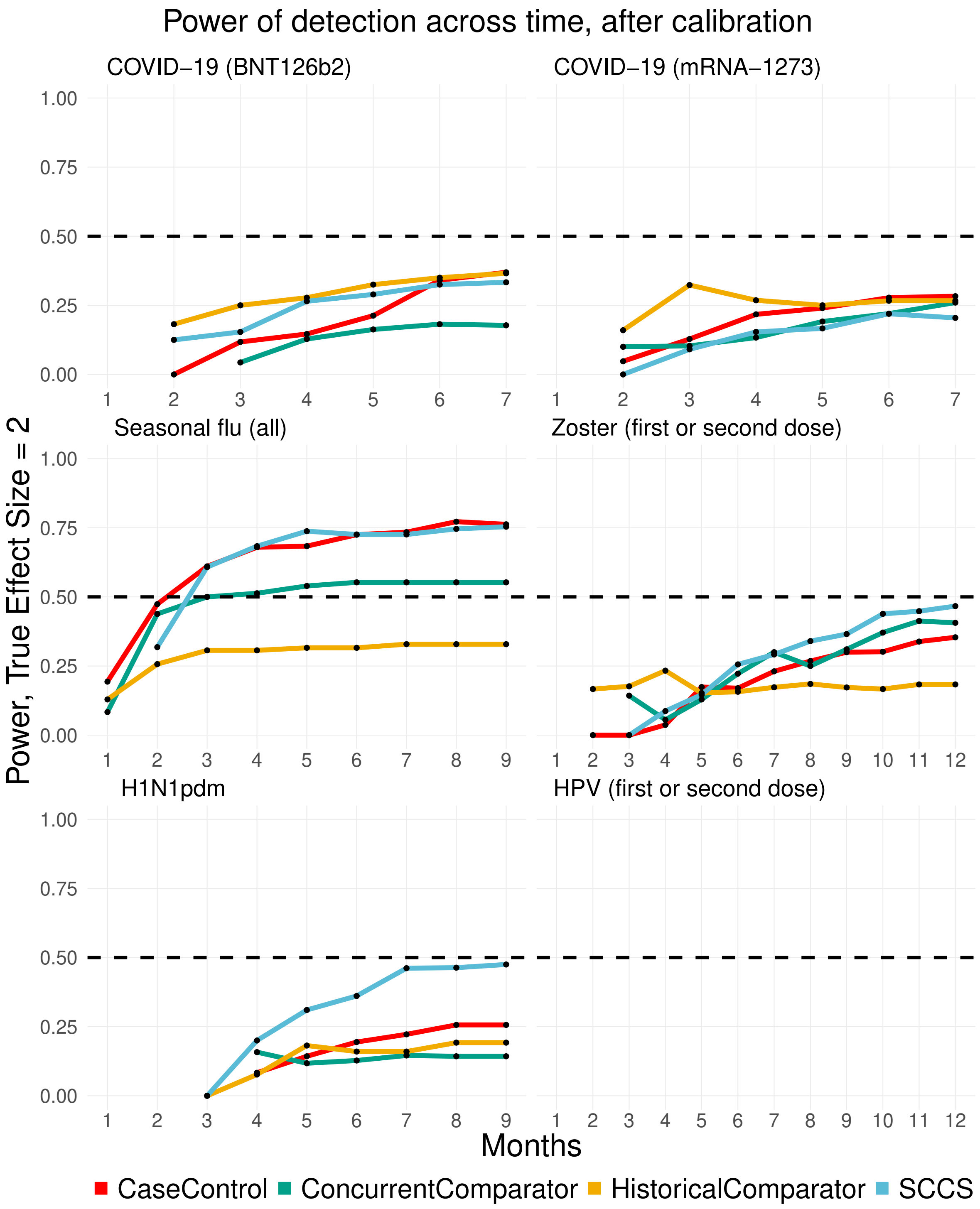


Figure 25: Power of detection (true effect size = 2) of the methods as a function of time, across different exposures for the MDCR data source.

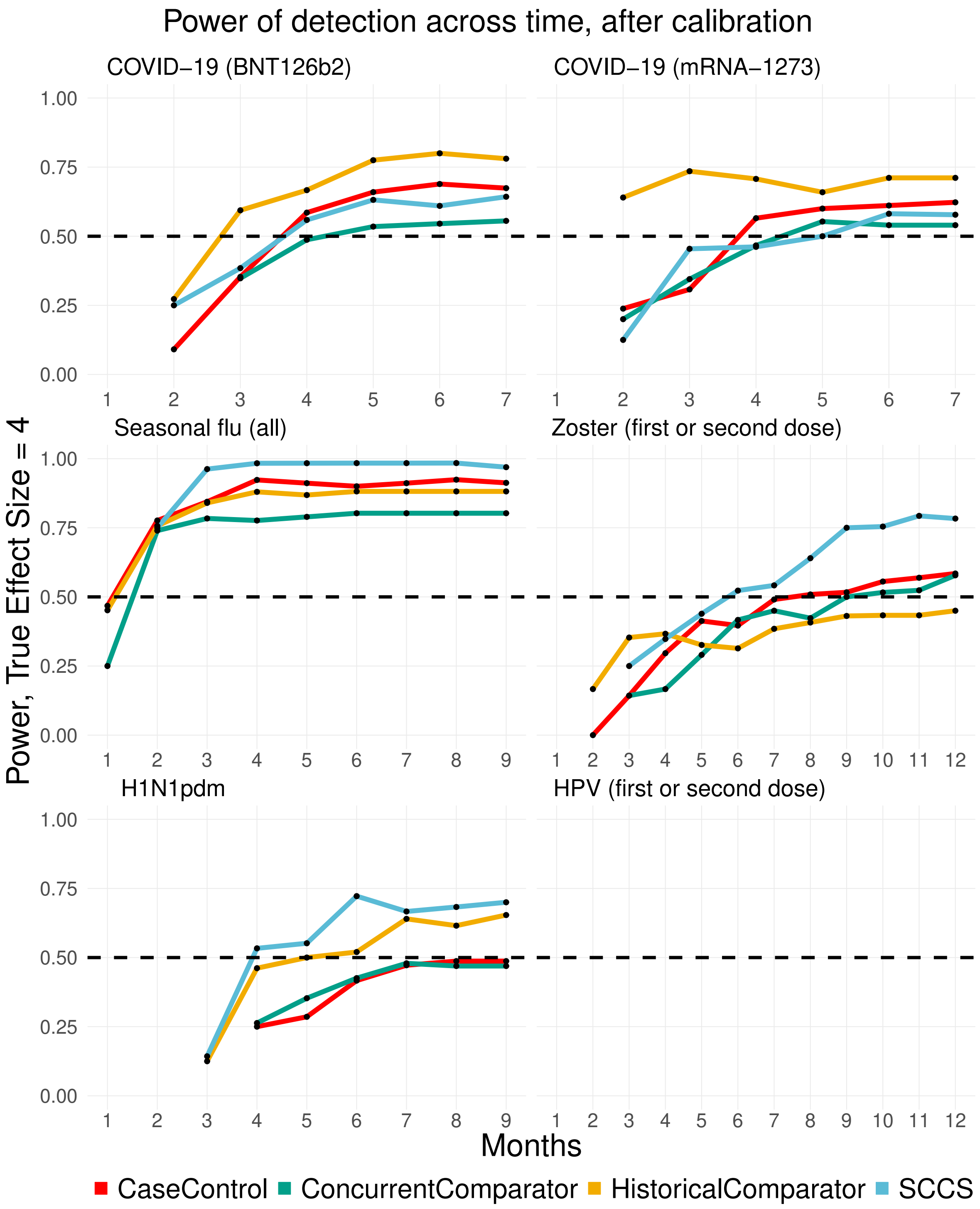


Figure 26: Power of detection (true effect size = 4) of the methods as a function of time, across different exposures for the MDCR data source.

### 3 Proportion of non-finite estimates

Table 1: Proportion of non-finite estimates.

| | Data Source | | | | |
|---|---|---|---|---|---|
| | MDCR | MDCD | Optum EHR | Optum DOD | CCAE |
| **COVID-19 vaccination (BNT126b2)** | | | | | |
| Concurrent comparator | 0.52 | 0.23 | 0.02 | 0.19 | 0.16 |
| SCCS | 0.23 | 0.22 | 0.26 | 0.27 | 0.14 |
| Historical comparator | 0.47 | 0.20 | 0.02 | 0.17 | 0.15 |
| Case control | 0.46 | 0.20 | 0.04 | 0.17 | 0.17 |
| **COVID-19 vaccination (mRNA-1273)** | | | | | |
| Concurrent comparator | 0.46 | 0.22 | 0.08 | 0.16 | 0.19 |
| SCCS | 0.16 | 0.19 | 0.23 | 0.24 | 0.18 |
| Historical comparator | 0.40 | 0.18 | 0.06 | 0.17 | 0.16 |
| Case control | 0.41 | 0.19 | 0.09 | 0.18 | 0.16 |
| **H1N1pdm vaccination** | | | | | |
| Concurrent comparator | 0.47 | 0.34 | 0.37 | 0.25 | 0.23 |
| SCCS | 0.02 | 0.00 | 0.01 | 0.01 | 0.01 |
| Historical comparator | 0.54 | 0.33 | 0.27 | 0.22 | 0.19 |
| Case control | 0.09 | 0.05 | 0.16 | 0.00 | 0.02 |
| **Seasonal flu vaccination (all)** | | | | | |
| Concurrent comparator | 0.18 | 0.18 | 0.01 | 0.15 | 0.15 |
| SCCS | 0.00 | 0.01 | 0.07 | 0.13 | 0.06 |
| Historical comparator | 0.14 | 0.13 | 0.00 | 0.10 | 0.10 |
| Case control | 0.00 | 0.00 | 0.01 | 0.00 | 0.00 |
| **HPV vaccination (first or second dose)** | | | | | |
| Concurrent comparator | – | 0.53 | 0.44 | 0.49 | 0.43 |
| SCCS | – | 0.00 | 0.00 | 0.00 | 0.00 |
| Historical comparator | – | 0.46 | 0.38 | 0.48 | 0.38 |
| Case control | – | 0.08 | 0.15 | 0.12 | 0.08 |
| **Zoster vaccination (first or second dose)** | | | | | |
| Concurrent comparator | 0.31 | 0.49 | 0.10 | 0.18 | 0.26 |
| SCCS | 0.00 | 0.00 | 0.00 | 0.04 | 0.01 |
| Historical comparator | 0.30 | 0.48 | 0.11 | 0.13 | 0.18 |
| Case control | 0.00 | 0.12 | 0.04 | 0.00 | 0.01 |

### 4 Results for the risk of myocarditis or pericarditis

#### 4.1 Results for the MDCR data source

Table 2: Risk of developing myocarditis or pericarditis following vaccination with COVID-19 BNT126b2 and mRNA-1273 vaccines for the MDCR data source. Each table shows the estimated risk ratio with the lower and upper bounds of the 95% confidence interval after empirical calibration, and whether or not a signal was declared.

| | Estimates | | |
|---|---|---|---|
| | Risk Ratio | 95% CI | Signal? |
| **COVID-19 vaccination (BNT126b2)** | | | |
| Concurrent comparator | 0.35 | 0.02 – 5.47 | No |
| SCCS | 0.78 | 0.15 – 4.00 | No |
| Historical comparator | 0.61 | 0.13 – 2.85 | No |
| Case control | 0.61 | 0.12 – 3.11 | No |
| **COVID-19 vaccination (mRNA-1273)** | | | |
| Concurrent comparator | 0.57 | 0.04 – 8.85 | No |
| SCCS | 1.38 | 0.21 – 8.81 | No |
| Historical comparator | 0.74 | 0.15 – 3.60 | No |
| Case control | 0.91 | 0.15 – 5.36 | No |

#### 4.2 Results for the MDCD data source

Table 3: Risk of developing myocarditis or pericarditis following vaccination with COVID-19 BNT126b2 and mRNA-1273 vaccines for the MDCD data source. Each table shows the estimated risk ratio with the lower and upper bounds of the 95% confidence interval after empirical calibration, and whether or not a signal was declared.

| | Estimates | | |
|---|---|---|---|
| | Risk Ratio | 95% CI | Signal? |
| **COVID-19 vaccination (BNT126b2)** | | | |
| Concurrent comparator | 1.18 | 0.47 – 2.94 | No |
| SCCS | 1.18 | 0.68 – 2.03 | No |
| Historical comparator | 1.40 | 0.72 – 2.73 | No |
| Case control | 1.55 | 0.80 – 3.01 | No |
| **COVID-19 vaccination (mRNA-1273)** | | | |
| Concurrent comparator | 1.07 | 0.46 – 2.50 | No |
| SCCS | 1.21 | 0.75 – 1.98 | No |
| Historical comparator | 1.23 | 0.67 – 2.28 | No |
| Case control | 0.92 | 0.49 – 1.72 | No |

#### 4.3 Results for the Optum DOD data source

Table 4: Risk of developing myocarditis or pericarditis following vaccination with COVID-19 BNT126b2 and mRNA-1273 vaccines for the Optum DOD data source. Each table shows the estimated risk ratio with the lower and upper bounds of the 95% confidence interval after empirical calibration, and whether or not a signal was declared.

| | **Estimates** | | |
|---|---|---|---|
| | Risk Ratio | 95% CI | Signal? |
| **COVID-19 vaccination (BNT126b2)** | | | |
| Concurrent comparator | 2.05 | 1.30 – 3.21 | Yes |
| SCCS | 1.50 | 1.13 – 1.99 | Yes |
| Historical comparator | 1.92 | 0.91 – 4.06 | No |
| Case control | 1.52 | 0.83 – 2.77 | No |
| **COVID-19 vaccination (mRNA-1273)** | | | |
| Concurrent comparator | 1.08 | 0.65 – 1.80 | No |
| SCCS | | | |
| Historical comparator | 1.26 | 0.60 – 2.64 | No |
| Case control | 1.20 | 0.62 – 2.33 | No |

#### 4.4 Results for the CCAE data source

Table 5: Risk of developing myocarditis or pericarditis following vaccination with COVID-19 BNT126b2 and mRNA-1273 vaccines for the CCAE data source. Each table shows the estimated risk ratio with the lower and upper bounds of the 95% confidence interval after empirical calibration, and whether or not a signal was declared.

| | **Estimates** | | |
|---|---|---|---|
| | Risk Ratio | 95% CI | Signal? |
| **COVID-19 vaccination (BNT126b2)** | | | |
| Concurrent comparator | 1.56 | 1.08 – 2.25 | Yes |
| SCCS | 1.23 | 1.00 – 1.50 | Yes |
| Historical comparator | 1.62 | 0.86 – 3.05 | No |
| Case control | 1.36 | 0.97 – 1.90 | No |
| **COVID-19 vaccination (mRNA-1273)** | | | |
| Concurrent comparator | 1.36 | 0.85 – 2.18 | No |
| SCCS | 1.49 | 1.18 – 1.89 | Yes |
| Historical comparator | 1.26 | 0.69 – 2.29 | No |
| Case control | 1.54 | 1.11 – 2.14 | Yes |
